# Time to Eat: Effects of Personalized Regular Meal Schedules on Body Weight and Mental and Physical Well-Being—A Randomized Controlled Pilot Trial

**DOI:** 10.1101/2024.01.30.24301983

**Authors:** Isabell Wilming, Jana Tuschewski, Jessie M. Osterhaus, Theresa J. G. Bringmann, Anisja Hühne-Landgraf, Olga Pivovarova-Ramich, Dominic Landgraf

## Abstract

**Background:** Circadian disruption is associated with metabolic and mental health disturbances, but whether increasing meal regularity can reduce body weight and improve well-being without prescribed dietary restriction is unclear.

**Objective:** To test the effects of personalized regular meal schedules on body weight and well-being in adults with body mass index (BMI) ≥22 kg/m².

**Methods:** In this single-center randomized controlled pilot trial, 121 adults were assigned 2:1 to personalized meal schedules derived from 2 weeks of app-based dietary records or to a minimal-intervention control using a self-selected 18-hour eating window. The intervention lasted 6 weeks. One hundred participants completed the trial and were included in the primary analysis. Models adjusted for baseline BMI, age, sex, and, for non-weight outcomes, the corresponding baseline value.

**Results:** Compared with controls, the intervention group had greater reductions in body weight (adjusted difference, −2.10 kg; 95% CI: −2.86, −1.34; P < 0.0001) and BMI (−0.70 kg/m²; 95% CI: −0.94, −0.45; P < 0.0001). Meal timing variability decreased substantially (−2.72 points; 95% CI: −3.13, −2.30; P < 0.0001), and greater improvements in regularity were associated with greater weight loss. Weight change was not associated with self-reported changes in energy intake or macronutrient composition. Among well-being outcomes, sleep quality showed the clearest adjusted between-group improvement (Pittsburgh Sleep Quality Index: −1.47 points; 95% CI: −2.28, −0.65; P = 0.0006).

**Conclusions:** Personalized regular meal schedules reduced body weight and were accompanied by improvements in well-being, most clearly sleep quality. Larger trials using objective metabolic and circadian measures are needed to confirm clinical utility and clarify mechanisms.

**Trial registration:** German Clinical Trials Register, DRKS00021419, registered August 21, 2020; https://drks.de/search/en/trial/DRKS00021419.

## 1. Introduction

Circadian clocks generate endogenous 24-hour rhythms across tissues, coordinating physiological processes with the external day–night cycle.^1,2^ This temporal organization relies on regular environmental time cues (*Zeitgebers*), among which food intake represents a particularly potent signal for peripheral metabolic regulation. Circadian disruption has been associated with adverse metabolic outcomes, including weight gain, as well as impairments in mental health.^3–14^

Interventions targeting the timing of food intake have gained increasing attention in humans. Several studies have demonstrated that restricting food intake to defined time periods can mitigate some of these adverse metabolic and mental effects.^15–19^ Time-restricted eating (TRE), which confines daily food intake to a limited time window, has been shown to induce modest weight loss, typically in the range of ∼0.9–3.5 kg over 12 weeks.^19–22^ Evidence further suggests that earlier eating windows may be metabolically advantageous.^23,24^ However, existing approaches define relatively broad time windows and do not explicitly address the regularity of eating behavior within these intervals.

Beyond restricting the time span of food intake, increasing the regularity of meals may represent an additional and largely unexplored behavioral dimension. From a circadian perspective, more stable temporal patterns of food intake could support a more consistent alignment between metabolic processes and recurring daily events. At the same time, such an approach does not require changes in caloric intake or food composition, potentially increasing feasibility and adherence under real-world conditions.

In the present randomized controlled pilot study, we hypothesized that increasing meal regularity through an individually derived eating schedule would reduce body weight and BMI and improve well-being in adults across a broad body mass index (BMI) range. To this end, participants’ habitual eating patterns were first characterized using a smartphone-based assessment, capturing individual temporal preferences, and individualized meal schedules were subsequently implemented in the experimental group (EG) for six weeks, while participants of the control group (CG) were instructed to confine their food intake to a self-selected 18-hour daily time window without further constraints, thereby largely maintaining their habitual eating patterns **(Fig. 1A)**.

**Figure 1.**
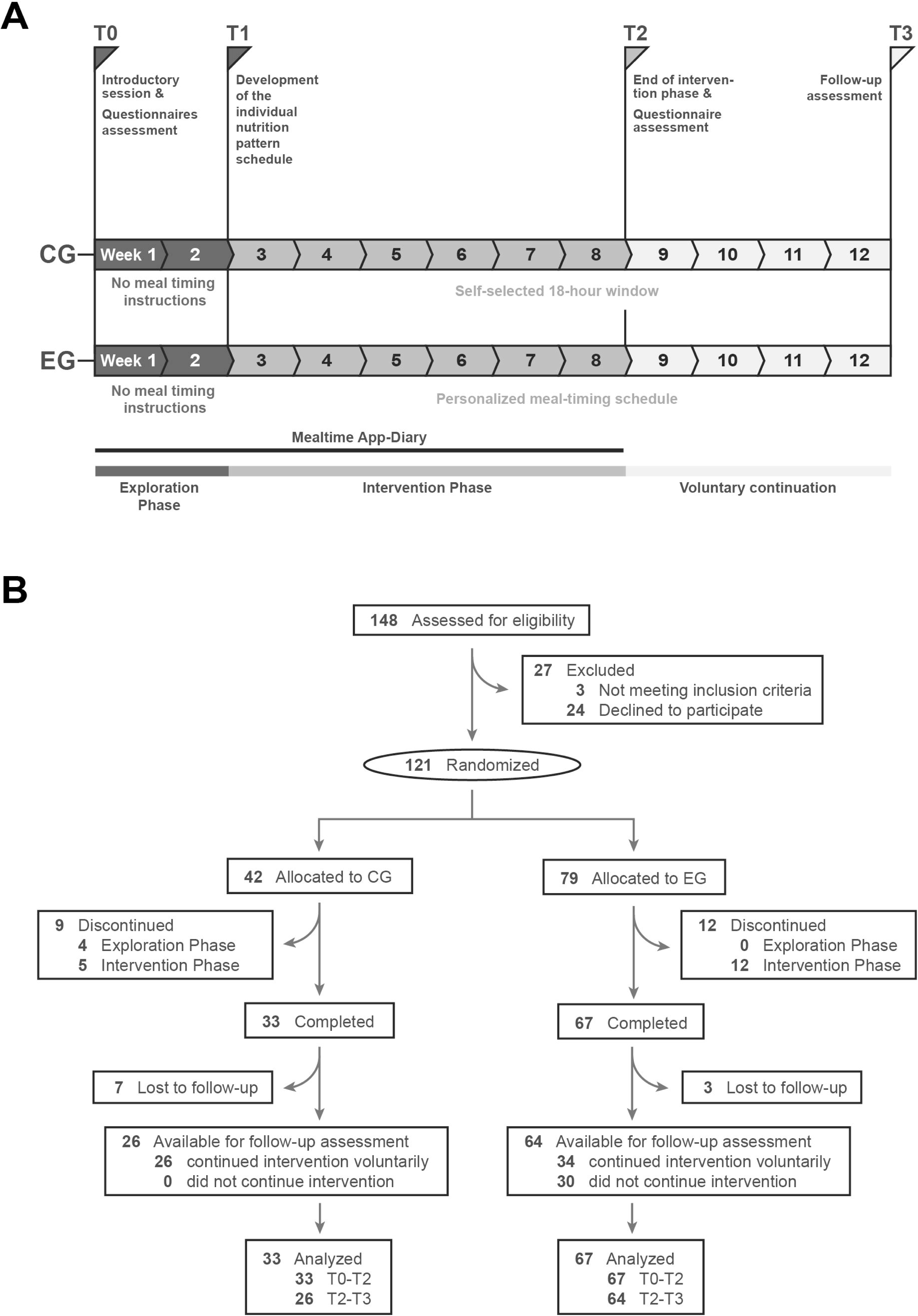
Study design and participant flow. (A) Timeline of the randomized controlled pilot study. During the exploration phase, randomized participants used the mealtime app-diary without receiving meal-timing instructions. After completion of the exploration phase, participants in the control group (CG) followed a self-selected 18-h eating window, whereas participants in the experimental group (EG) received an individualized meal-timing schedule based on their nutrition pattern. After completion of the intervention phase, participants in the EG could voluntarily continue the intervention until follow-up assessment. (B) Participant flow from eligibility assessment to analysis. If individual data were missing or excluded from specific analyses, the corresponding sample size and reason are provided in the respective figure legend.

The primary outcomes were changes in body weight and BMI, and secondary outcomes included multiple dimensions of physical and mental well-being. In addition, we examined whether changes in meal regularity were associated with weight loss and whether weight loss was related to self-reported caloric intake and diet composition. Further exploratory analyses were conducted to identify potential behavioral and temporal factors that may modulate the intervention effect, including baseline characteristics and individual temporal patterns.

Since the intervention focused on eating times without limiting calories or specific types of food, we termed this approach “TIME TO EAT”.

## 2. Methods

### Participants and ethics

This single-center, parallel-group randomized controlled pilot trial with a 2:1 allocation ratio used an exploratory framework to investigate the effects of increased meal timing regularity on body weight, BMI, and parameters related to self-reported well-being in adult participants. The study was conducted at the Clinic for Psychiatry and Psychotherapy of Ludwig Maximilian University Munich, Germany, and was prospectively registered at the German Clinical Trial Register (DRKS00021419). The study was approved by the Ethics Committee of Ludwig Maximilian University Munich under file number 19-975. All participants provided informed consent before participation. The study protocol is available from the corresponding author upon reasonable request. No separate statistical analysis plan was prospectively prepared. Patients and members of the public were not involved in the design, conduct, or reporting of the trial.

Participants were recruited across Germany between September 2020 and August 2021, with final follow-up assessments completed between October and November 2021. Recruitment was conducted through flyers at universities, fitness studios, adult education centers, pharmacies, and grocery stores, as well as through social media. Inclusion criteria were age between 18 and 65 years, body mass index (BMI) of at least 22 kg/m², and the ability to understand the study procedures. Exclusion criteria included concurrent dieting, regular use of medication affecting appetite or body weight, diagnosed metabolic, psychiatric, or addictive disorders, pregnancy, blindness, bedriddenness, or dependence on assistance with eating.

A total of 148 participants were recruited, of whom 121 were randomized to the experimental group (EG; n = 79) or control group (CG; n = 42). One hundred participants completed the intervention and were included in the primary analysis (EG: n = 67; CG: n = 33). Sex was recorded as self-reported male or female and is reported in Table S1. Gender identity, ancestry, race, ethnicity, and socioeconomic status were not systematically collected. The absence of these data limits assessment of generalizability across demographic and socioeconomic groups.

Participants were randomized at baseline (T0) in a 2:1 ratio to the experimental or control group. The random allocation sequence was generated by IW and JMO using the randomization function in Microsoft Excel. Simple randomization without blocking or stratification was used, with allocation probabilities corresponding to the intended 2:1 ratio. The allocation sequence was accessible to the study personnel responsible for enrollment and assignment and was therefore not concealed. IW, JT, JMO, and TJGB enrolled participants, assigned them to the interventions, and had access to the random allocation sequence.

Participants were blinded to the existence of an alternative study condition and were therefore unaware of their comparative group assignment. They received information and instructions only for their assigned condition. Study personnel delivering the interventions and supervising body weight measurements were not blinded to group assignment. Data analysts were also not blinded to group assignment.

### Study design

Each participant completed a 12-week program consisting of baseline assessment, a two-week exploration phase, a six-week intervention phase, and a four-week follow-up period (**Fig. 1A**). No concomitant care or lifestyle interventions beyond the assigned study procedures were prescribed or restricted during the trial. Due to COVID-19 restrictions, all study procedures, including instruction, monitoring, and data collection, were conducted remotely via video-based communication. Body weight and height were measured by participants under live supervision by study staff.

At baseline (T0), eligibility was confirmed, demographic data were collected, and participants received a standardized introduction to the study procedures. Body weight and height were measured under remote supervision. Participants then completed validated questionnaires assessing physical and mental well-being, sleep quality, self-efficacy, depressive symptoms, and chronotype (**Tab. S3**). At the end of the exploration phase (T1), body weight was reassessed under remote supervision. At the end of the intervention phase (T2), body weight was again reassessed under remote supervision, and participants completed the same questionnaires as at baseline. Four weeks after completion of the intervention (T3), participants were contacted via email to report current body weight and to indicate whether they had continued to follow the meal timing recommendations. No dietary records were collected during the follow-up period.

### Dietary recording

During the two-week exploration phase, participants were instructed to maintain their habitual eating behavior while documenting all caloric intake events, including meals, snacks, and caloric beverages, using a smartphone-based dietary tracking application (Fddb Extender, FDDB Internetportale GmbH, Berlin, Germany). The application recorded the timing, composition, and estimated quantity of food intake.^25,26^ Participants were instructed to document intake as accurately as possible, including portion sizes and additional descriptive details. Study staff had access to the recorded data to monitor compliance and download entries for analysis.

During the six-week intervention phase, participants in both groups continued to document all caloric intake using the smartphone application. Participants in the EG adhered to their individualized meal schedules, whereas participants in the CG maintained their habitual eating patterns within the specified time window. Body weight was self-measured weekly and reported to study staff. Participants were to be withdrawn from the study if their BMI decreased below 19 kg/m² during the intervention. This safety threshold was not reached by any participant. Adverse events were not otherwise systematically assessed. Compliance was supported through at least one mid-intervention contact and additional follow-up in cases of missing diary entries.

### Personalized meal schedules

Individualized meal schedules were generated for participants in the EG based on their recorded meal timing data from the exploration phase. For each participant, clusters of eating times were identified using a data-driven approach combining the elbow method to determine the optimal number of clusters and k-means clustering to estimate representative meal times. Cluster centers were calculated as mean values and subsequently rounded to practical time intervals. Proposed meal times were discussed with participants and adjusted if necessary to accommodate individual constraints such as work schedules. Participants were then instructed to consume meals only at these predefined times, allowing a tolerance window of ±30 min. Missed meals were not to be compensated outside scheduled time windows.

Participants in the CG underwent a similar review of their baseline eating behavior but were not provided with structured meal timing. Instead, they were instructed to confine food intake to a self-selected 18-h daily window without further restrictions, serving as a minimal-intervention control condition. ^18^ Participants in the CG additionally received sham psychoeducation on intermittent fasting that did not address circadian rhythms or meal regularity.

### Outcome measures

The prespecified co-primary outcomes were changes in body weight and BMI from the end of the exploration phase (T1) to the end of the six-week intervention phase (T2). Secondary outcomes included changes from baseline (T0) to the end of the intervention phase (T2) in physical and mental well-being, sleep quality, depressive symptoms, self-efficacy, and chronotype. Group-level outcomes were summarized as mean changes, and adjusted between-group differences in change were estimated as described in the Statistical analysis section. Self-reported well-being was assessed using validated questionnaires, including the 36-item Short Form Health Survey (SF-36)^27^, the Pittsburgh Sleep Quality Index (PSQI)^28^, the Self-Assessment Inventory of Depressive Symptoms (IDS-SR)^29^, and the Scale of General Self-Efficacy (GSE)^30^ (**Table S3**). For visualization purposes, composite scores for physical and mental health were derived by averaging z-standardized SF-36 subscale scores across participants and time points. Chronotype was assessed using the Munich Chronotype Questionnaire (MCTQ)^31^ and represented by the mid-sleep phase on free days corrected for sleep debt (MSFsc). Eating behavior was continuously assessed through real-time smartphone-based documentation, allowing extraction of meal timing, energy intake, and macronutrient composition.

### Mealtime variability score

Meal timing regularity was quantified using a Mealtime Variability Score (MTVS), developed for this study as a categorical measure of temporal deviation between actual eating events and predefined meal times. Each intake event was assigned a score based on its deviation from the closest scheduled meal time: 1 = +/− 0-15 min, 2 = +/− 16-30 min, 3 = +/− 31-45 min, 4 = +/− 46-60 min, 5 = +/− 61-75 min, 6 = +/− 76-90 min, 7 = +/− 91-105 min, 8 = +/− 106-120 min, 9 = +/− 2-3 h, 10 = +/− 3-4 h, 11 = over +/− 4 h.^32^ Lower scores indicate higher regularity. Average MTVS values were calculated across meals and time periods to derive individual measures of eating regularity.

### Changes to the trial protocol

No changes were made to the prespecified primary outcomes or intervention conditions after trial commencement. Three prespecified secondary measures were not implemented as originally planned. The Hamilton Depression Rating Scale (HAMD) was not administered because COVID-19-related remote study procedures precluded the face-to-face assessment considered necessary for reliable administration. The Positive and Negative Affect Schedule (PANAS) was briefly piloted in a small subset of participants as a potential alternative but was discontinued because it was considered insufficiently comprehensive for the intended assessment. A study-specific questionnaire was not implemented because it had not been formally validated. Additional analyses of meal-timing characteristics and potential predictors of intervention response were conducted post hoc and are reported as exploratory analyses.

### Statistical analysis

The primary analysis population for body weight and BMI comprised the 100 participants who completed the intervention and had measurements available at T1 and T2. Participants were analyzed according to their randomized group assignment. The primary analyses of body weight and BMI were complete-case analyses. Missing values in other longitudinal datasets were handled using mixed-effects models with restricted maximum likelihood estimation where applicable. Sensitivity analyses for body weight and BMI additionally included randomized participants with available baseline data using last observation carried forward, as described below.

Statistical analyses were performed using SPSS version 24, Python, GraphPad Prism version 9.2.0, and R. Longitudinal group differences and time effects were analyzed using repeated-measures two-way ANOVA for complete datasets. For datasets containing missing values, mixed-effects models using restricted maximum likelihood estimation were applied instead, as implemented in GraphPad Prism. Bonferroni correction was used for multiple comparisons. Within-group comparisons between exploration and intervention phases were assessed using paired t tests. In cases involving unequal sample sizes, Welch’s correction was applied.

Associations between variables were assessed using linear regression analyses. For the co-primary outcomes of body weight and BMI, the primary analyses compared changes from the exploration phase (T1) to the end of the intervention phase (T2) between the randomized groups using multiple linear regression models adjusted for baseline BMI, age, and sex. For dietary, meal timing, and well-being outcomes, adjusted between-group differences in change were estimated using multiple linear regression models additionally adjusted for the baseline value of the respective outcome. Adjusted between-group differences are reported as regression coefficients (β) with 95% confidence intervals. Within-group changes shown in the tables represent unadjusted mean changes with 95% confidence intervals. Cohen’s d was calculated from unadjusted change scores as a standardized between-group effect size.

A post hoc sensitivity analysis for body weight and BMI included randomized participants with available baseline data using last observation carried forward for drop-outs. Last observation carried forward was defined as the last actual available post-baseline value carried forward; if a weekly post-T1 body weight value was available, this value was used rather than automatically carrying forward the T1 value.

Exploratory multivariable regression analyses were performed in the EG to assess independent predictors of BMI reduction. Predictors included change in meal regularity, change in energy intake, change in eating interval, change in sleep–eating phase alignment, and weekday–weekend meal timing discrepancy. Multicollinearity was assessed using variance inflation factors. Residual normality was assessed using normality tests implemented in GraphPad Prism.

The exact number of participants included in each analysis is reported in the corresponding figure legends, tables, or results sections. Unless otherwise stated, values are reported as mean ± SD or mean with 95% confidence intervals, as indicated in the respective tables and figure legends. All statistical tests were 2-sided, and P values <0.05 were considered statistically significant.

Sample size was estimated a priori for this randomized pilot study based on expected body weight loss derived from a previous time-restricted eating study, as no prior study using personalized scheduled meal times was available. In that study, 10-hour time-restricted eating over 12 weeks was associated with a mean body weight reduction of 3.3 kg and an SD of 3.2 kg. For the present study, we assumed an expected body weight reduction of approximately 2.5 kg and a large standardized effect size of Cohen’s d = 0.8, which was converted to f = 0.4 for the power analysis. The significance level was set at α = 0.05. An a priori power analysis was conducted in G*Power using the F-test family and a repeated-measures design comprising 2 groups and 3 assessment time points (T0, T1, and T2), resulting in a target sample size of 100 participants. No interim analyses were planned or conducted, and no trial-level stopping guidelines were specified.

## 3. Results

### 3.1. Participant characteristics

A total of 148 participants were recruited for this pilot study, of whom 121 were randomized to the EG (n = 79) or CG (n = 42) **(Fig. 1B)**. One hundred participants completed the intervention (EG: n = 67; CG: n = 33). Drop-outs were primarily due to illness/accident or constraints that made it impossible to adhere to fixed mealtimes. No participant reached the predefined BMI safety threshold of <19 kg/m².

At baseline, participants in both groups were, on average, in the overweight range. Mean BMI was 27.4 ± 4.6 kg/m² in the EG and 25.5 ± 3.8 kg/m² in the CG, and mean age was 38.2 ± 14.4 and 32.6 ± 11.8 years, respectively (**Table S1**). Participants in the EG were also slightly older than those in the CG (38.2 ± 14.4 vs. 32.6 ± 11.8 years; p = 0.0440). There were no significant differences in sex distribution or baseline dietary intake between groups. Baseline BMI did not differ between completers and drop-outs in either group (CG: p = 0.2207; EG: p = 0.4437; **Fig. S1A**).

### 3.2. Intervention fidelity: increased meal regularity

Study participants documented all caloric intake events, including meals, snacks, and caloric beverages, using a smartphone-based dietary tracking application during a two-week exploration and a six-week intervention phase. In total, 13,838 and 34,564 caloric intake events from completers were analyzed during the exploration and intervention phases, respectively. Based on data-driven clustering of individual eating-time patterns, most EG participants were assigned to three individualized meal-time clusters (n = 53), while some showed four clusters (n = 13), and one participant showed two clusters, which subsequently formed the basis for the personalized meal schedules.

Meal timing variability was assessed using a Mealtime Variability Score (MTVS), a categorical measure in which higher values indicate greater irregularity (see Methods for definition). At baseline, both groups showed meal irregularity, with MTVS values around 4, corresponding to an average deviation of approximately ±45–60 minutes per meal.

During the intervention, meal regularity improved substantially in the EG, with MTVS values decreasing from 3.879 (95% CI [3.658, 4.101]) to 1.723 (95% CI [1.573, 1.873]) (within-group p < 0.0001; **Fig. 2A-D**), corresponding to a deviation lower than ±16–30 minutes per meal. In contrast, no improvement was observed in the CG; instead, a slight increase in variability was noted (from 3.353 [95% CI 3.066, 3.640] to 3.936 [95% CI 3.681, 4.192]; within-group p = 0.0011).

**Fig. 2.**
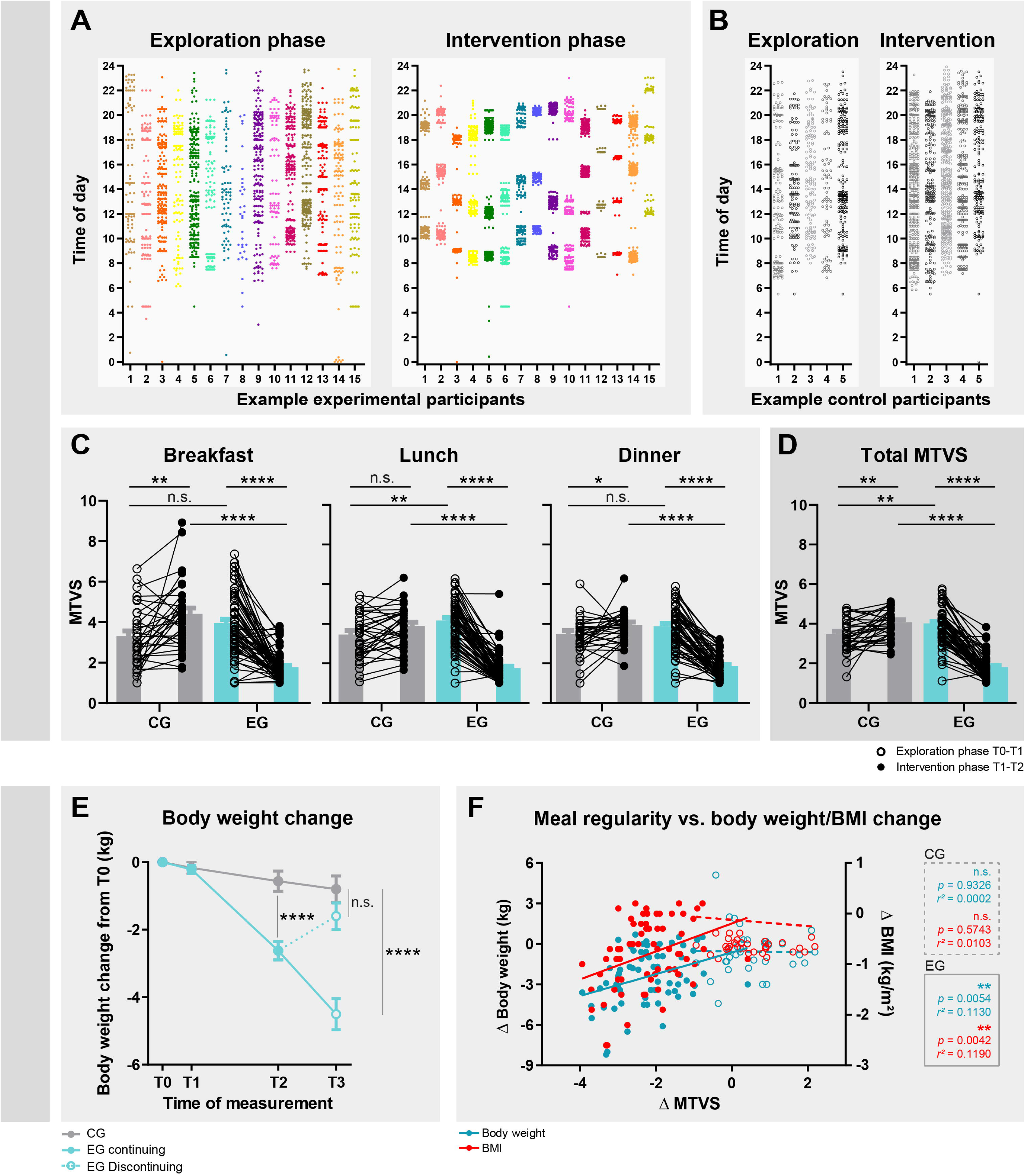
Increased meal regularity, body weight changes, and their association. (A, B) Timing of caloric intake events in example participants of the experimental group (EG) (A) and control group (CG) (B) during exploration and intervention. Each point represents one caloric intake event. (C, D) Changes in mealtime variability score (MTVS) for breakfast, lunch, and dinner (C) and across all caloric intake events (D). Lower MTVS values indicate greater meal regularity. (E) Body weight change from baseline (T0 = 0); EG participants are stratified during follow-up by continuation or discontinuation of the intervention. (F) Associations between changes in MTVS and changes in body weight or BMI. Statistical annotations indicate repeated-measures ANOVA or mixed-effects models with Bonferroni correction and linear regression analyses where applicable. Adjusted between-group analyses are reported in the main text and tables.

Between-group comparisons confirmed a significant reduction in meal timing variability in the EG compared to the CG (interaction p < 0.0001), indicating successful implementation of the intervention.

### 3.3. Body weight and BMI changes

During the baseline phase, no significant changes in body weight or BMI were observed in either group. After six weeks of intervention, EG participants showed significantly greater reductions in body weight and BMI compared to CG participants after adjustment for baseline BMI, age, and sex (body weight: β = −2.10 kg, 95% CI [−2.86, −1.34], p < 0.0001; BMI: β = −0.70 kg/m², 95% CI [−0.94, −0.45], p < 0.0001) (**Figs. 2E**, **Table 1**). Individual trajectories further illustrated substantial variability in weight-loss responses, including pronounced reductions in some EG participants maintaining the intervention (**Fig. S1B,C**). From baseline assessment at T0 to the end of the intervention at T2, participants in the EG lost on average 2.62 kg (95% CI [−3.16, −2.08], p < 0.0001) and 0.87 kg/m² (95% CI [−1.05, −0.69], p < 0.0001). In contrast, changes in body weight and BMI from T0 to T2 were not significant in the CG (body weight: −0.56 kg, p = 0.0918; BMI: −0.20 kg/m², p = 0.0658).

**Table 1:** Changes in anthropometric, dietary, and meal-timing outcomes from the exploration phase (T1) to the end of the intervention phase (T2), by randomized group.

| Outcome | n (CG/EG)<sup>c</sup> | T2-T1 change in CG<sup>a</sup> |  | T2-T1 change in EG<sup>a</sup> |  | Adjusted difference between groups<sup>b</sup> |  |  |
| --- | --- | --- | --- | --- | --- | --- | --- | --- |
|  |  | Mean (95% CI) | P value | Mean (95% CI) | P value | β (95% CI) | P value | Cohen's d<sup>e</sup> |
| Body weight, kg | 33/67 | -0.3909 (-0.9125 to 0.1307) | 0.1367 | -2.41 (-2.906 to -1.915) | <0.0001 | -2.095 (-2.855 to -1.335) | <0.0001 | 1.08 |
| BMI, kg/m<sup>2</sup> | 33/67 | -0.1400 (-0.3104 to 0.03036) | 0.1039 | -0.8076 (-0.9717 to -0.6436) | <0.0001 | -0.6951 (-0.9449 to -0.4453) | <0.0001 | 1.08 |
| MTVS Total<sup>d</sup> | 33/67 | 0.5833 (0.2624 to 0.9043) | 0.0008 | -2.179 (-2.412 to -1.946) | <0.0001 | -2.716 (-3.130 to -2.302) | <0.0001 | 2.94 |
| Energy Intake, kJ<sup>d</sup> | 30/67 | 36.25 (-371.2 to 443.7) | 0.8569 | -347.6 (-627.6 to -67.67) | 0.0157 | -494.5 (-976.4 to -12.61) | 0.0444 | 0.34 |
| Fat, (g/1,000 kJ)<sup>d</sup> | 30/67 | 0.4920 (0.07340 to 0.9106) | 0.0228 | -0.1873 (-0.5708 to 0.1962) | 0.333 | -0.4799 (-0.9330 to -0.02683) | 0.0381 | 0.47 |
| Carbohydrate, (g/1,000 kJ)<sup>d</sup> | 30/67 | -0.7803 (-1.743 to 0.1827) | 0.1083 | -0.02478 (-0.6979 to 0.6484) | 0.9416 | 0.6562 (-0.3632 to 1.676) | 0.2042 | 0.28 |
| Protein, (g/1,000 kJ)<sup>d</sup> | 30/67 | -0.2877 (-0.7824 to 0.2071) | 0.244 | 0.4164 (0.08984 to 0.7430) | 0.0132 | 0.6779 (0.1453 to 1.210) | 0.0132 | 0.53 |
| Eating Interval, h<sup>d</sup> | 32/67 | 0.4406 (-0.3111 to 1.192) | 0.241 | -2.008 (-2.515 to -1.501) | <0.0001 | -2.295 (-3.051 to -1.540) | <0.0001 | 1.18 |
| Breakfast time, h<sup>d</sup> | 33/67 | -0.3336 (-0.7535 to 0.08619) | 0.1153 | 1.128 (0.7943 to 1.462) | <0.0001 | 1.287 (0.7709 to 1.804) | <0.0001 | 1.11 |
| Dinner time, h<sup>d</sup> | 32/67 | 0.2659 (-0.04278 to 0.5747) | 0.0888 | -0.6607 (-0.9294 to -0.3921) | <0.0001 | -0.9408 (-1.368 to -0.5138) | <0.0001 | 0.90 |

<sup>a</sup> Within-group changes were unadjusted.
<sup>b</sup> Between-group differences were adjusted for baseline BMI, sex, and age.
<sup>c</sup> Values indicate the numbers of participants with data available at both relevant time points and included in the corresponding change analysis, shown as CG/EG.
<sup>d</sup> For dietary and meal timing outcomes, models were additionally adjusted for the baseline value of the respective outcome.
<sup>e</sup> Cohen's d was calculated from unadjusted change scores.

The findings were similar in a post hoc sensitivity analysis in which missing post-baseline values for participants who discontinued the study were handled using last observation carried forward. After adjustment for baseline BMI, age, and sex, the intervention group continued to show significantly greater reductions in body weight (β = −1.62 kg, 95% CI [−2.32, −0.92], p < 0.0001) and BMI (β = −0.53 kg/m², 95% CI [−0.77, −0.30], p < 0.0001) compared to the CG.

At the four-week follow-up, 34 participants in the EG reported maintaining regular meal timing, whereas others reported returning to more irregular eating patterns. Participants who reported maintaining regularity continued to lose body weight (mean −1.12 kg, p < 0.0001), whereas those reporting a return to irregular eating showed a weight increase (mean +0.48 kg, p = 0.0016). Accordingly, a similar pattern was observed for BMI (continuing: −0.36 kg/m², p < 0.0001; discontinuing: +0.17 kg/m², p = 0.0018). As dietary intake and meal timing were no longer recorded during this phase and classification was based solely on participant self-report, these follow-up findings should be considered exploratory.

Within the EG, further exploratory analyses indicated that higher baseline BMI was associated with greater BMI reduction, although the explained variance was small (R² = 0.0597, p = 0.0465).

### 3.4. Association between meal regularity and weight loss

To examine the extent to which weight loss in the EG was related to improvements in meal timing regularity, we correlated changes in body weight and BMI with individual changes in meal regularity (ΔMTVS).

Consistent with our hypothesis, a significant association was observed between improved meal regularity and weight loss in the EG (body weight: R² = 0.1130, p = 0.0054; BMI: R² = 0.1190, p = 0.0042), whereas no such relationship was found in the CG (body weight: R² = 0.0002345, p = 0.9326; BMI: R² = 0.01029, p = 0.5743) (**Fig. 2F**).

A breakdown by individual meals showed that improvements in the regularity of lunch and dinner were significantly associated with reductions in body weight (lunch: R² = 0.1192, p = 0.0054; dinner: R² = 0.1159, p = 0.0048) and BMI (lunch: R² = 0.1142, p = 0.0052; dinner: R² = 0.1257, p = 0.0032) (**Fig. S2**). No significant association was observed for breakfast.

### 3.5. Relation between weight change and self-reported food quantity and composition

To assess whether the observed weight loss in the EG could be related to changes in dietary intake, self-reported energy intake and macronutrient composition were analyzed.

Average daily energy intake did not differ significantly between groups during the exploration phase (**Table S1**). Repeated-measures analyses showed no significant time × group interaction for self-reported energy intake. Within the EG, a small reduction in absolute self-reported energy intake was observed (from 7071 to 6723 kJ/day; p = 0.0157), and adjusted analyses indicated a modest between-group difference in energy intake change (β = −494.5 kJ/day, 95% CI [−976.4, −12.61], p = 0.0444) (**Fig. 3A**). Cumulative self-reported energy intake patterns remained stable across study phases, arguing against progressively changing reporting behavior over time (**Fig. S3A,B**).

**Fig. 3.**
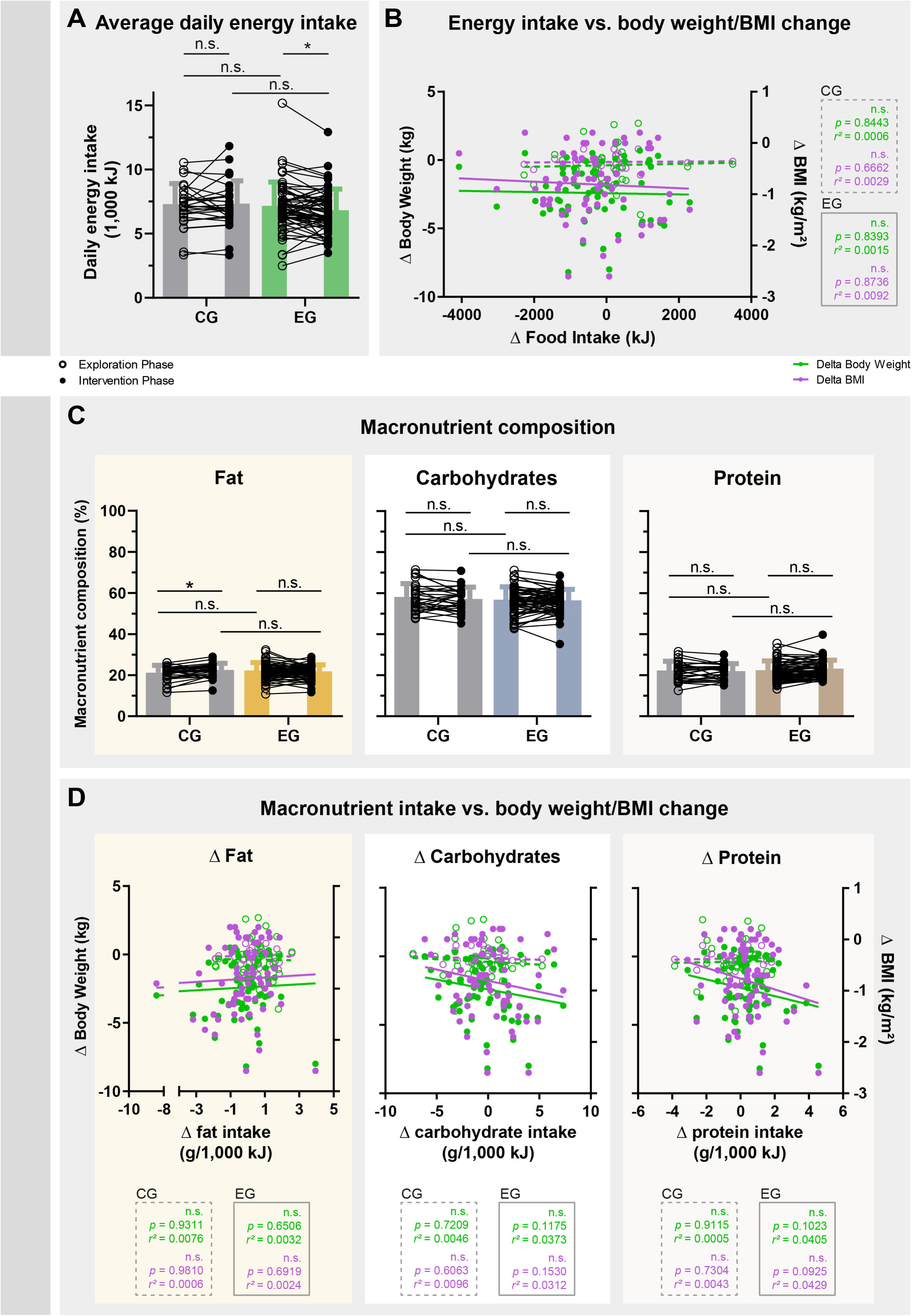
Self-reported energy intake and macronutrient composition showed no consistent association with weight loss. (A) Average daily energy intake during the exploration and intervention phases in the control group (CG) and experimental group (EG). (B) Associations between changes in energy intake and changes in body weight or BMI. (C) Macronutrient composition during exploration and intervention, shown as percentage of total intake from fat, carbohydrates, and protein. (D) Associations between changes in macronutrient intake and changes in body weight or BMI. Statistical annotations indicate repeated-measures ANOVA or mixed-effects models with Bonferroni correction and linear regression analyses where applicable. Adjusted between-group analyses are reported in the main text and tables.

At the individual level, participants reported heterogeneous changes in intake, with some indicating increased and others decreased consumption (**Fig. 3A**). However, self-reported changes in energy intake were not associated with changes in body weight or BMI (**Fig. 3B**).

Macronutrient composition remained broadly stable, although small adjusted between-group differences were observed for fat and protein intake (**Figs. 3C**). Repeated-measures analyses revealed no significant time × group interactions for carbohydrate or protein intake, whereas a small interaction effect was observed for fat intake (p = 0.0095). Consistent with this, adjusted analyses of changes from exploration to intervention showed no significant between-group difference for carbohydrate intake, whereas only small group differences were observed for fat and protein intake. Importantly, self-reported changes in macronutrient composition were not associated with changes in body weight or BMI (all p ≥ 0.09) (**Fig. 3D**).

### 3.6. Exploratory analyses of additional meal timing-related associations with weight loss

To further characterize factors potentially associated with the intervention effect, a series of exploratory analyses was performed. These analyses were not pre-specified as primary outcomes and should be interpreted as hypothesis-generating.

#### 3.6.1. Additional meal timing characteristics

Besides meal regularity, the intervention affected additional aspects of eating behavior, including eating interval and meal timing (**Fig. 4A**). Repeated-measures analyses showed a significant reduction of the eating interval in the EG compared to the CG (time × group interaction p < 0.0001). In addition, shifts in meal timing were observed, with later first meals and earlier last meals in the EG. Adjusted analyses accounting for baseline values, baseline BMI, age, and sex confirmed significant between-group differences in changes of eating interval as well as in the timing of the first and last meals. However, changes in eating interval as well as in the timing of the first and last meals were not associated with weight loss (**Figs. 4B–D**).

**Fig. 4.**
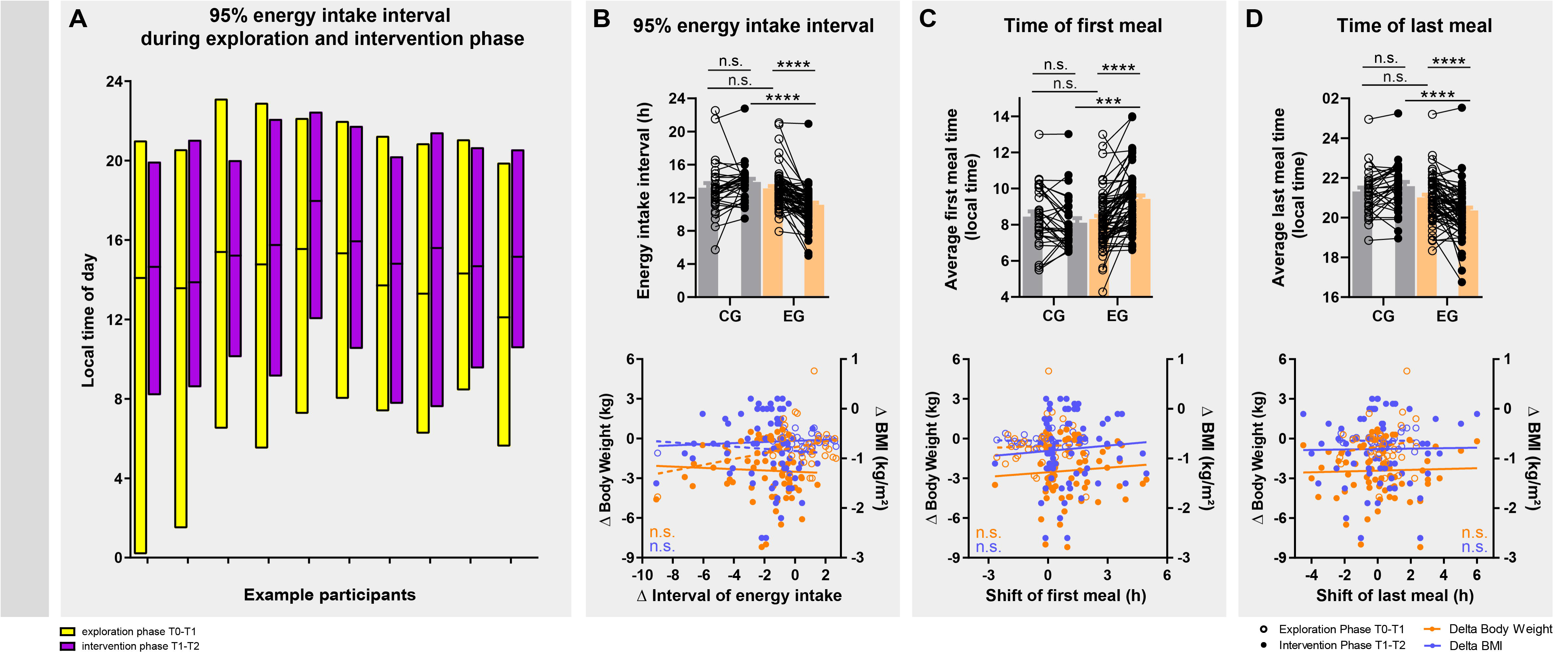
Additional meal timing characteristics and their association with weight loss. (A) Daily 95% caloric intake interval in 10 example participants during the exploration and intervention phases. Bars indicate the time window containing 95% of caloric intake events. (B) Changes in eating interval duration and their associations with changes in body weight or BMI. (C, D) Changes in the timing of the first (C) and last (D) caloric intake event and their associations with changes in body weight or BMI. Statistical annotations indicate repeated-measures ANOVA or mixed-effects models with Bonferroni correction and linear regression analyses where applicable. Adjusted between-group analyses are reported in the main text and tables.

The intervention further influenced the consistency of meal timing across the week. Separate clustering of weekday and weekend eating patterns showed reduced variability for breakfast- and lunch-associated meal clusters, whereas dinner timing remained largely unchanged (**Fig. S4A**). However, despite the absence of a significant group-level effect, individual reductions in weekday–weekend variability in dinner timing (“dinner jet lag”) were associated with greater reductions in BMI (**Fig. S4B**).

#### 3.6.2. Personalization of meal timing and its association with weight loss

To explore whether alignment of meal timing with individual temporal preference is associated with intervention success, we analyzed the relationship between chronotype and eating timing. Chronotype was represented by the mid-sleep phase on free days corrected for sleep debt (MSFsc), reflecting the midpoint of the individual sleep period while accounting for differences in sleep duration between workdays and free days.^31^ To characterize meal timing, an analogous measure, the mid-eat phase (mEP), was calculated as the midpoint between the first and last caloric intake of the day.

A significant association between MSFsc and mEP was observed during the exploration phase (R² = 0.2084, p = 0.0001), indicating that habitual eating timing reflects individual temporal preference (**Fig. S4C,D**). During the intervention, this association became stronger (R² = 0.3286, p < 0.0001), suggesting a more consistent alignment between sleep timing and meal timing.

To quantify this alignment at the individual level, residuals from the MSFsc –mEP relationship were calculated. Exploratory analyses indicated that a closer alignment between these measures was associated with greater reductions in BMI (Δ residuals vs. Δ BMI: R² = 0.1084, p = 0.0074) (**Fig. S4E**).

#### 3.6.3. Multivariable analysis of meal-related predictors

A multivariable regression model including meal regularity, caloric intake, eating interval, and timing-related variables was used in EG to assess independent predictors of BMI reduction.

In this model, improved meal regularity remained an independent predictor (estimate = 0.2582, p = 0.0185). Caloric intake and eating interval were not significant contributors. Reduction in weekday–weekend differences in dinner timing was independently associated with BMI reduction (estimate = −0.3071, p = 0.0008) (**Table S2**).

### 3.7. Well-being outcomes

Several measures of physical and mental well-being improved over the course of the intervention in the EG, whereas no consistent improvements were observed in the CG.

Sleep quality improved significantly in the EG, as assessed by the Pittsburgh Sleep Quality Index^28^ (PSQI; within-group p < 0.0001), while no significant change was observed in the CG (**Fig. 5A**; **Table 2**). Similarly, depressive symptoms decreased significantly in the EG (IDS-SR^29^; within-group p < 0.0001), and self-efficacy increased significantly over time (GSE^30^; within-group p < 0.0001). In contrast, the CG showed no comparable pattern of improvement across these measures.

**Fig. 5.**
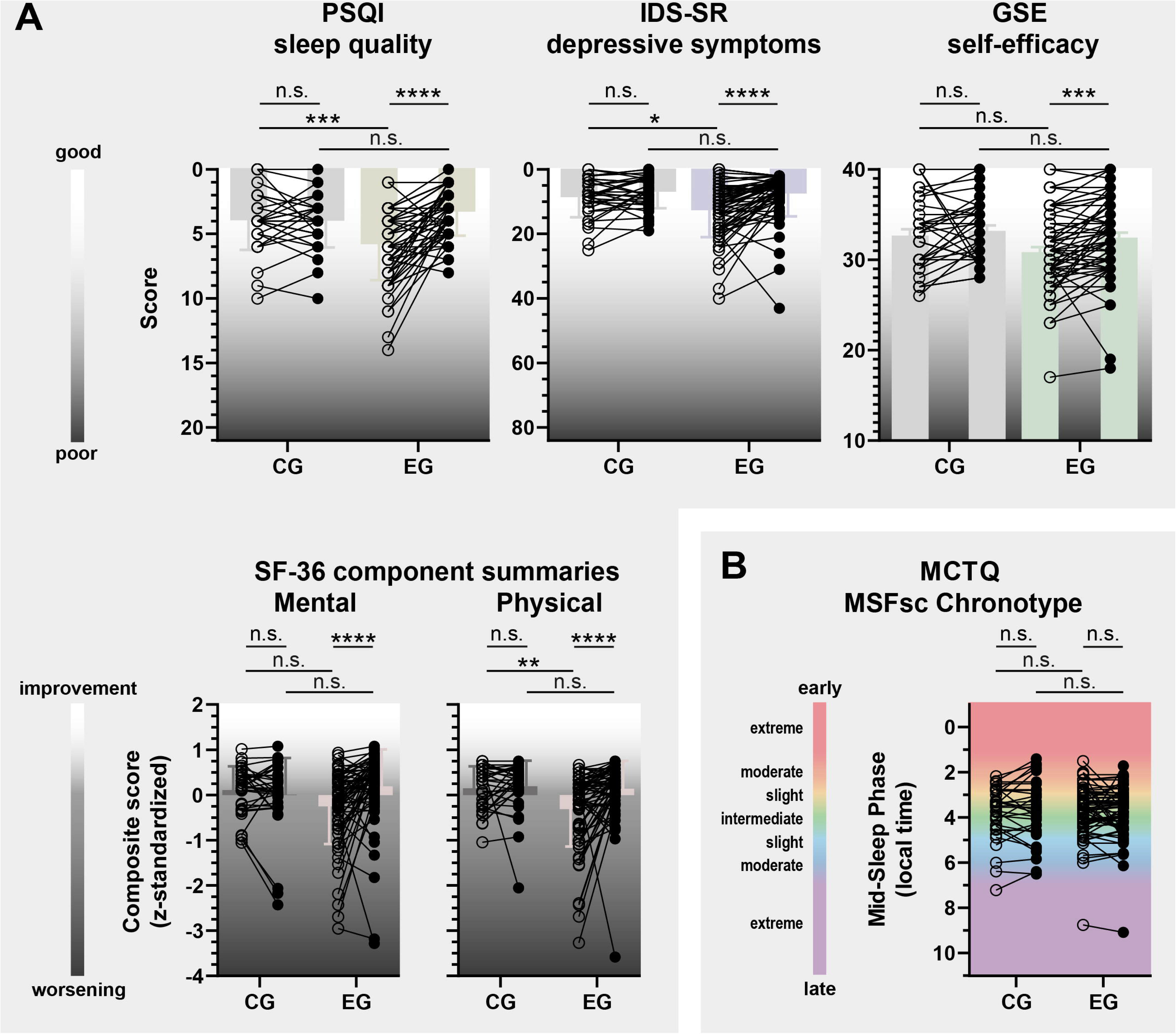
Changes in self-reported well-being and chronotype during the intervention. (A) Sleep quality (PSQI), depressive symptoms (IDS-SR), self-efficacy (GSE), and SF-36 mental and physical component summary scores (MCS and PCS) in the control group (CG, gray) and experimental group (EG, colored). Lower PSQI and IDS-SR scores and higher GSE and SF-36 scores indicate improvement. SF-36 component scores were derived from z-standardized subscale scores. (B) Chronotype, assessed as MSFsc, remained stable in both groups. Statistical annotations indicate repeated-measures ANOVA or mixed-effects models with Bonferroni correction. Adjusted between-group analyses are reported in Table 2.

**Table 2:**
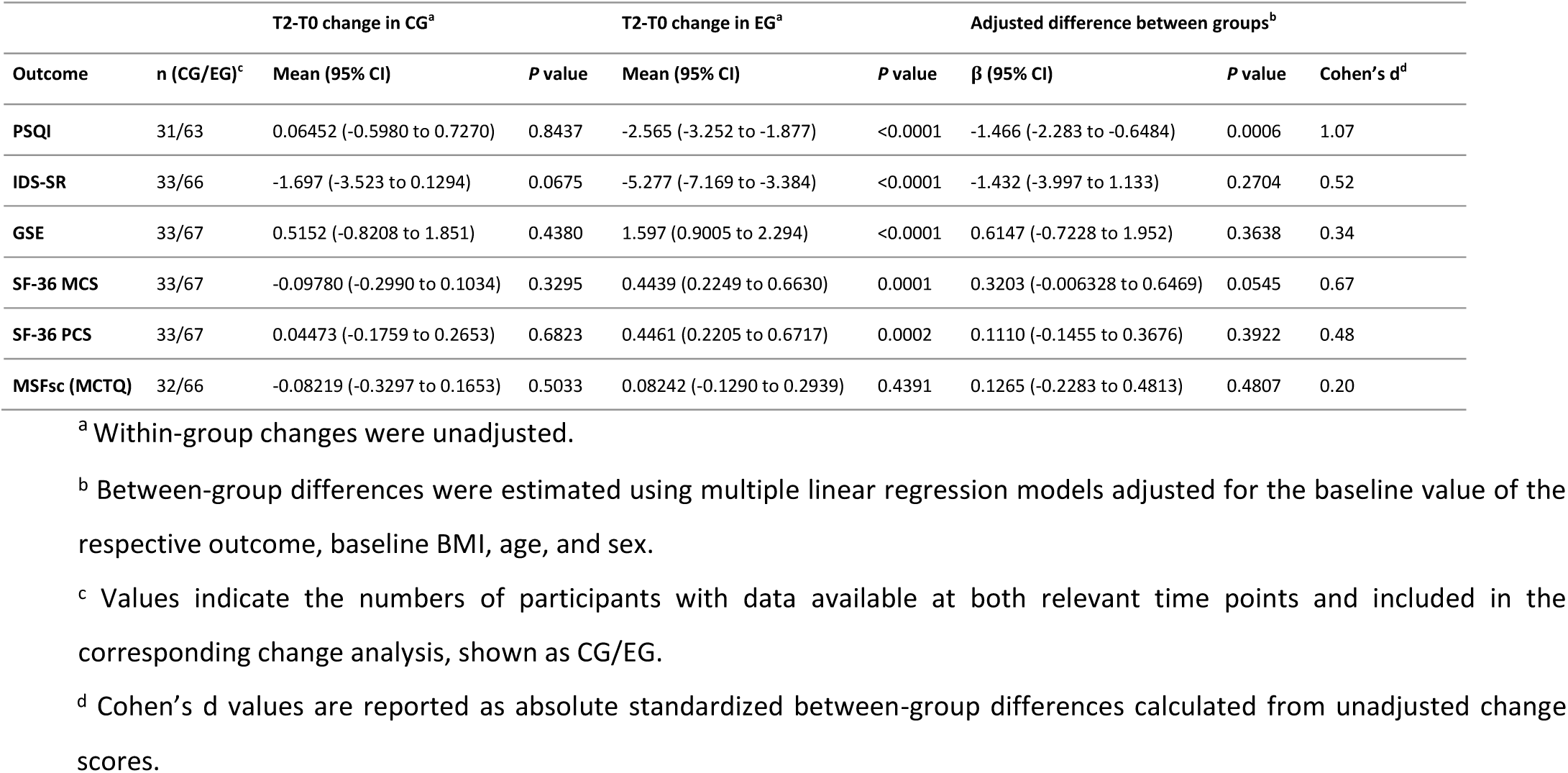
Changes in well-being and chronotype outcomes from baseline (T0) to the end of the intervention phase (T2), by randomized group.

Consistent improvements were also observed for SF-36-derived well-being measures^27^. In the EG, both the mental and physical SF-36 component summary scores increased significantly from exploration to intervention (MCS: within-group p = 0.0001; PCS: within-group p = 0.0002), indicating improved self-reported mental and physical health status (**Fig. 5A**; **Table 2**). Analyses of individual SF-36 domains further showed significant improvements in several subscales, including physical functioning, general health, and vitality (**Fig. S5**).

Adjusted between-group analyses supported these findings most clearly for sleep quality (PSQI: β = −1.47, 95% CI [−2.28, −0.65], p = 0.0006). Improvements in the SF-36 mental component summary score showed a similar direction but did not reach statistical significance (MCS: β = 0.32, 95% CI [−0.006, 0.65], p = 0.0545). No significant adjusted between-group effects were observed for the SF-36 physical component summary score, depressive symptoms, self-efficacy, or chronotype measures (**Table 2**). Thus, the intervention was associated with broad within-group improvements in self-reported well-being in the EG, with the most robust adjusted between-group evidence observed for sleep quality.

The chronotype marker MSFsc derived from the MCTQ^31^ did not change over the course of the study in either group (adjusted between-group difference: p = 0.4807), consistent with the notion that the intervention stabilized behavioral patterns without shifting underlying temporal preference (**Fig. 5B**; **Table 2**).

## 4. Discussion

In this randomized pilot study, increasing meal regularity resulted in significant reductions in body weight and BMI without prescribed changes in energy intake or diet composition. Importantly, weight loss was associated with improvements in meal timing regularity, but not with self-reported changes in energy intake or macronutrient composition.

Interventions targeting the timing of food intake have gained increasing attention in recent years, particularly in the context of TRE. TRE protocols typically confine food intake to broad daily time windows and have been associated with modest weight loss, often requiring longer intervention periods or additional dietary constraints.^19–23,26,33,34^ In contrast, the present study specifically targeted the regularity of eating behavior within the day, rather than the duration or timing of an overall eating window. Although our intervention also led to secondary changes in eating behavior, such as a shortening of the daily eating interval or avoiding late meals, these parameters were not associated with weight loss, suggesting that increased temporal regularity represents a distinct and potentially more specific behavioral component.

Notably, weight loss was not related to self-reported changes in energy intake or macronutrient composition. A small reduction in self-reported energy intake of approximately 350 kJ per day was observed in the EG, and adjusted analyses indicated a modest between-group difference in energy intake change. Similar spontaneous reductions in energy intake have also been reported in other meal timing-based interventions, such as time-restricted eating, and may contribute to their beneficial metabolic effects.^35^ However, changes in energy intake were not associated with weight loss in the present study. Exploratory analyses of macronutrient composition indicated that carbohydrate intake remained stable across study phases, whereas only small differences were observed for fat and protein intake. Importantly, these changes were not associated with weight or BMI reduction, arguing against macronutrient composition as a primary driver of the observed effects. Although a contributory role of reduced energy intake cannot be fully excluded, the absence of any detectable association – despite considerable inter-individual variability – supports the interpretation that temporal aspects of eating behavior, rather than quantitative dietary changes, were linked to the observed weight loss.

The lack of a corresponding association in the CG further strengthens this interpretation. Although some variability in meal timing was observed among control participants, these changes were small compared to the intervention-induced improvements in regularity. This suggests that a certain magnitude and consistency of temporal regularity may be required to elicit measurable effects on body weight.

Adjusted analyses further demonstrated that the observed reductions in body weight and BMI remained significant after accounting for baseline BMI, age, and sex, supporting the robustness of the intervention effect despite baseline differences between groups.

Exploratory analyses provided additional insights into factors that may modulate the intervention effect. The broad BMI range was chosen to allow exploratory assessment of whether intervention response varied with baseline BMI. Higher baseline BMI was weakly associated with greater BMI reduction within the EG, providing preliminary support for this hypothesis. However, the small effect size and the absence of a formal group-by-baseline BMI interaction preclude conclusions about whether baseline BMI modifies the intervention effect. Furthermore, analyses of individual meals suggested that improvements in the regularity of lunch and dinner were more strongly associated with weight loss than those of breakfast. These findings should be interpreted cautiously, as they were not primary outcomes, but they may inform the design of future studies.

Additional exploratory analyses addressed the role of individual temporal patterns. Although the intervention did not explicitly aim to align eating times with circadian phase, habitual eating timing was associated with individual temporal preference, and improved alignment between sleep timing and meal timing was associated with greater reductions in BMI. These findings suggest that personalization of meal timing may contribute to intervention efficacy, although this hypothesis was not directly tested in the present study.

Previous experimental work in animals provides a potential framework for interpreting these findings. In mice, restricting food intake to the active phase prevents weight gain and metabolic dysfunction despite identical energy intake.^17,36^ These observations support the concept that temporal organization of feeding behavior can influence metabolic outcomes independently of energy intake.

Several mechanisms could potentially underlie the observed association between meal regularity and weight loss. Circadian systems are known to anticipate recurring environmental events such as food intake and to coordinate metabolic processes accordingly.^37–40^ Increased temporal regularity may therefore support this anticipatory regulation and could contribute to improved metabolic efficiency. Experimental studies in animals and observational data in humans further suggest that regular feeding patterns can influence endogenous circadian rhythms and support their stabilization.^23,37–41^ While such effects were not directly assessed in the present study, it remains possible that increased meal regularity contributed to a more consistent temporal organization of physiological processes. In addition, circadian regulation of the microbiome and of rhythmic gene expression in metabolic tissues may represent possible pathways through which meal timing regularity could influence energy utilization.^42–47^

Beyond these circadian-informed mechanisms, alternative or complementary explanations should be considered. First, despite real-time recording, unintentional misreporting of dietary intake cannot be fully excluded.^48^ Irregular eating patterns may be more prone to omissions (e.g., snacks or caloric beverages) and estimation errors, whereas more structured eating could facilitate more consistent reporting, potentially introducing subtle biases in estimated energy intake. However, even under the assumption that dietary intake was reported accurately, additional mechanisms remain plausible. Behavioral aspects of how food is consumed, rather than what is consumed, may contribute, as regular meal patterns could influence eating rate, portion structuring, or satiety signaling.^49^ Furthermore, more consistent meal timing may reduce metabolic variability across the day, potentially stabilizing postprandial responses, including diet-induced thermogenesis, substrate utilization, and insulin dynamics, thereby influencing energy storage efficiency.^11^ Regular eating patterns may also affect digestive processes and nutrient absorption, leading to small differences in effective energy availability. Finally, structured daily routines may indirectly influence energy expenditure in everyday life, for example through more consistent patterns of spontaneous physical activity, which were not assessed in the present study.

In addition to effects on body weight, the intervention was associated with improvements in several self-reported well-being measures within the EG, with the most robust between-group evidence observed for sleep quality. Improvements in mental well-being showed a similar direction, whereas adjusted between-group effects for broader physical and mental health measures were less consistent. While these outcomes were secondary and the study was not powered specifically for these measures, the overall pattern suggests that increased regularity of daily behaviors may extend beyond metabolic regulation.

One possible explanation is that more structured daily routines provide a stabilizing framework for both physiological and behavioral processes. Regular meal timing may contribute to a more predictable daily structure, potentially supporting sleep and mental well-being. Alternatively, improvements may reflect indirect effects of weight loss or increased perceived control over daily habits. As these mechanisms were not directly assessed, the observed associations should be interpreted cautiously and require confirmation in future studies.

Given the well-established links between metabolic function and mental health, these findings may be of particular relevance for future interventions targeting both domains.

### 4.1. Limitations

In our pilot study, we did not collect objective metabolic or circadian physiological parameters that could provide mechanistic explanations for the observed weight loss. Therefore, conclusions regarding underlying biological mechanisms remain speculative. While behavioral regularity of eating clearly increased, potential effects on endogenous physiological rhythms were not directly assessed and should be addressed in future studies using objective metabolic and circadian measurements.

Another limitation is the reliance on participant-measured body weight and self-reported eating behavior and dietary intake. Although body weight assessments were conducted under live video supervision and dietary intake was recorded in real time using a smartphone application validated for dietary compliance assessment^25^, measurement and reporting biases cannot be excluded^48^. However, cumulative energy intake patterns remained relatively stable across study phases, arguing against a marked phase-specific shift in reporting behavior.

The study was conducted under real-world conditions, enhancing ecological validity but limiting experimental control. In addition, the study design does not allow disentangling the specific contribution of personalization from the effects of increased meal regularity.

The relatively short intervention period and modest sample size limit conclusions regarding long-term effectiveness, and the primary complete-case analysis may have introduced attrition bias.

Given the number of secondary and exploratory analyses, these findings should be interpreted cautiously and as hypothesis-generating.

Finally, baseline BMI differed between groups after randomization of the final sample. However, adjusted analyses demonstrated that the observed intervention effects on body weight and BMI remained significant after accounting for baseline BMI, age, and sex, arguing against baseline differences as the primary explanation for the observed effects.

### 4.2. Conclusion

In summary, this randomized pilot study showed that increasing meal regularity reduced body weight over a relatively short period without prescribed changes in energy intake or diet composition and was accompanied by improvements in physical and mental well-being measures, with the clearest adjusted between-group evidence observed for sleep quality.

These findings support the concept that temporal organization of eating behavior represents a relevant and previously underappreciated behavioral dimension of metabolic regulation. Given the close link between metabolic and affective disturbances, such an approach may be particularly relevant in the context of common comorbid conditions.

Due to its simplicity and compatibility with everyday routines, this intervention may facilitate adherence compared to more restrictive dietary approaches, although this remains to be tested in future studies.

Overall, these findings provide a rationale for larger, confirmatory trials incorporating objective metabolic and circadian measurements to further evaluate the clinical potential of this approach.

## Data Availability

All data produced in the present study are available upon reasonable request to the authors. The codes used to calculate MTVS, individual optimal number of meals, and personalized optimal eating times will be available at https://github.com/dolandgraf/Time-To-Eat.git as of the date of publication of this paper.

## Disclaimer

None.

## Abbreviations

BMI: body mass index
CG: control group
EG: experimental group
GSE: General Self-Efficacy Scale
IDS-SR: Inventory of Depressive Symptomatology–Self-Report
MCS: mental component summary
MCTQ: Munich Chronotype Questionnaire
mEP: mid-eat phase
MSFsc: mid-sleep phase on free days corrected for sleep debt
MTVS: Mealtime Variability Score
PCS: physical component summary
PSQI: Pittsburgh Sleep Quality Index
SF-36: 36-Item Short Form Health Survey
TRE: time-restricted eating

## Acknowledgments

We thank all participants who took part in this study. We also thank Thomas Schneider-Axmann (Ludwig Maximilian University Munich, Germany) and Elisabeth Paul (Linköping University, Sweden) for valuable statistical advice.

## Author Contributions

DL, AHL, and IW designed the research; IW, JT, JMO, TJGB, and DL conducted the research; IW and JT developed the study software; DL analyzed the data and performed the statistical analyses; DL prepared the figures; DL and OPR wrote the original draft of the manuscript; DL, AHL, IW, and OPR critically reviewed and edited the manuscript; DL supervised the research; DL and OPR had primary responsibility for the final content; and all authors read and approved the final manuscript.

## Data sharing

De-identified individual participant data described in the manuscript and the corresponding codebook will be made available upon reasonable request to the corresponding author, pending approval of the proposed use and subject to applicable data-protection and ethical requirements. Available analytic code will be provided with the data. The custom code used to calculate the Mealtime Variability Score, determine the individualized number of meal-time clusters, and generate personalized meal schedules will be made publicly and freely available without restriction at https://github.com/dolandgraf/Time-To-Eat upon publication.

## Declaration of Generative AI and AI-assisted Technologies in the Writing Process

During the preparation and revision of this work, the corresponding author used ChatGPT (OpenAI) to assist with language editing, clarity, sentence structure, manuscript organization, and checking consistency with journal reporting and formatting requirements. The tool was not used to generate or analyze study data or to perform statistical analyses. All AI-assisted content was critically reviewed, verified, and edited by the authors, who take full responsibility for the content of the publication.

