## Supplementary material for "Time to Eat: Effects of Personalized Regular Meal Schedules on Body Weight and Mental and Physical Well-Being—A Randomized Controlled Pilot Trial": Legends of supplemental figures

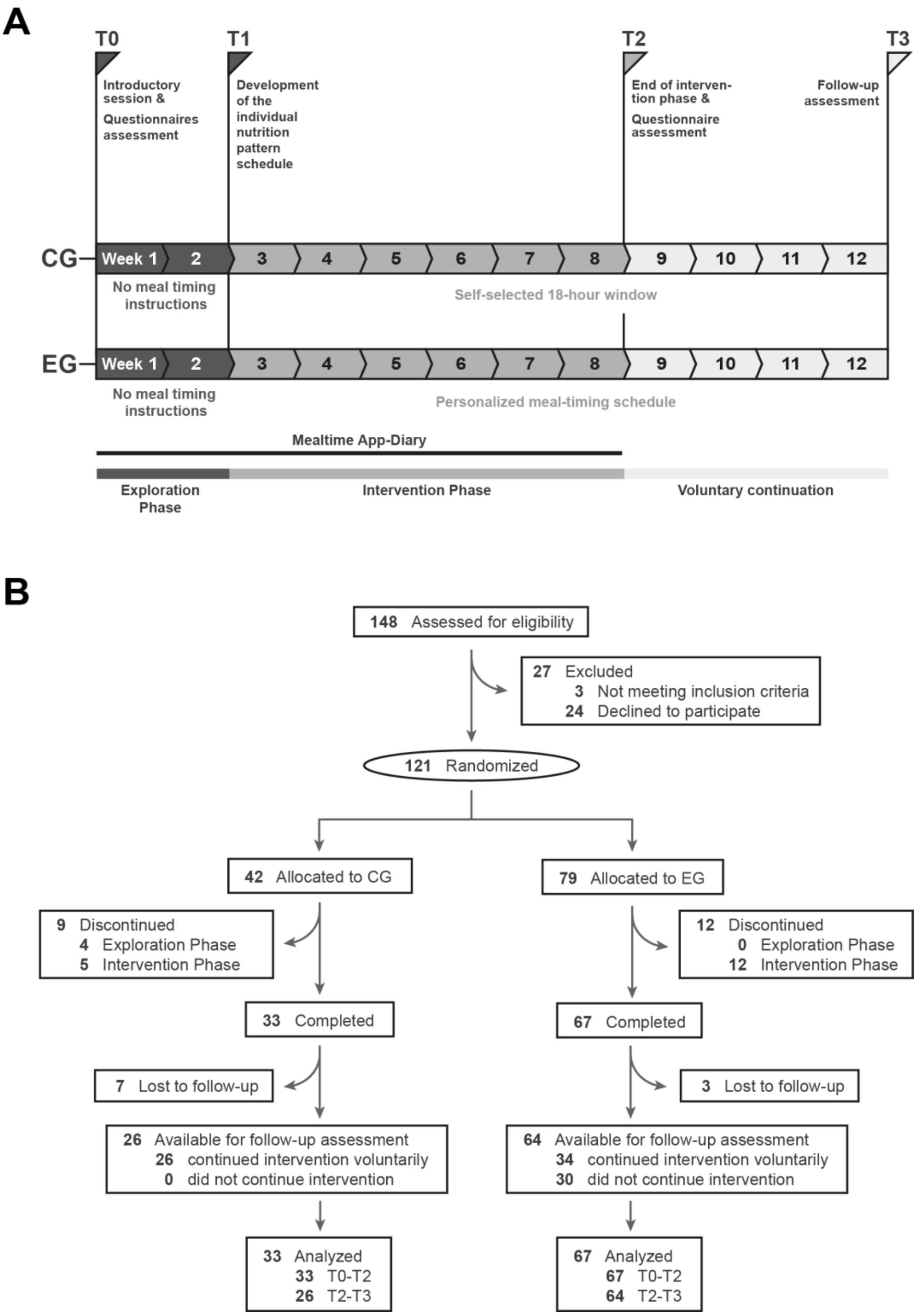


**Figure 1. Study design and participant flow.** (A) Timeline of the randomized controlled pilot study. During the exploration phase, participants used the mealtime app-diary without receiving meal-timing instructions. After randomization, participants in the control group (CG) followed a self-selected 18-h eating window, whereas participants in the experimental group (EG) received an individualized meal-timing schedule based on their nutrition pattern. After completion of the intervention phase, participants in the EG could voluntarily continue the intervention until follow-up assessment. (B) Participant flow from eligibility assessment to analysis. If individual data were missing or excluded from specific analyses, the corresponding sample size and reason are provided in the respective figure legend.

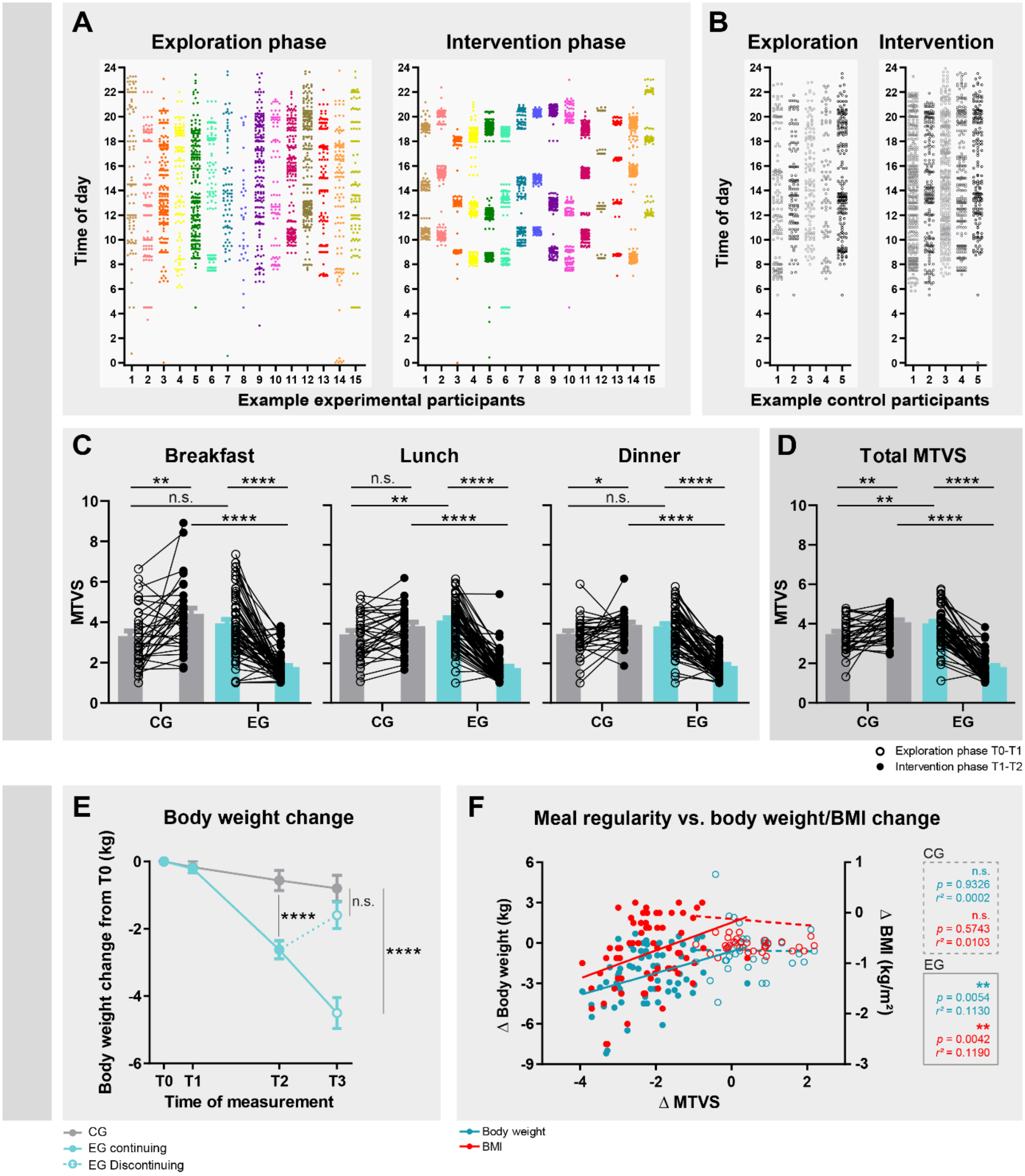


Fig. 2. Increased meal regularity, body weight changes, and their association. (A, B) Timing of caloric intake events in example participants of the experimental group (EG) (A) and control group (CG) (B) during exploration and intervention. Each point represents one caloric intake event. (C, D) Changes in mealtime variability score (MTVS) for breakfast, lunch, and dinner (C) and across all caloric intake events (D). Lower MTVS values indicate greater meal regularity. (E) Body weight change from baseline (T0 = 0); EG participants are stratified during follow-up by continuation or discontinuation of the intervention. (F) Associations between changes in MTVS and changes in body weight or BMI. Statistical annotations indicate repeated-measures ANOVA or mixed-effects models with Bonferroni correction and linear regression analyses where applicable. Adjusted between-group analyses are reported in the main text and tables.

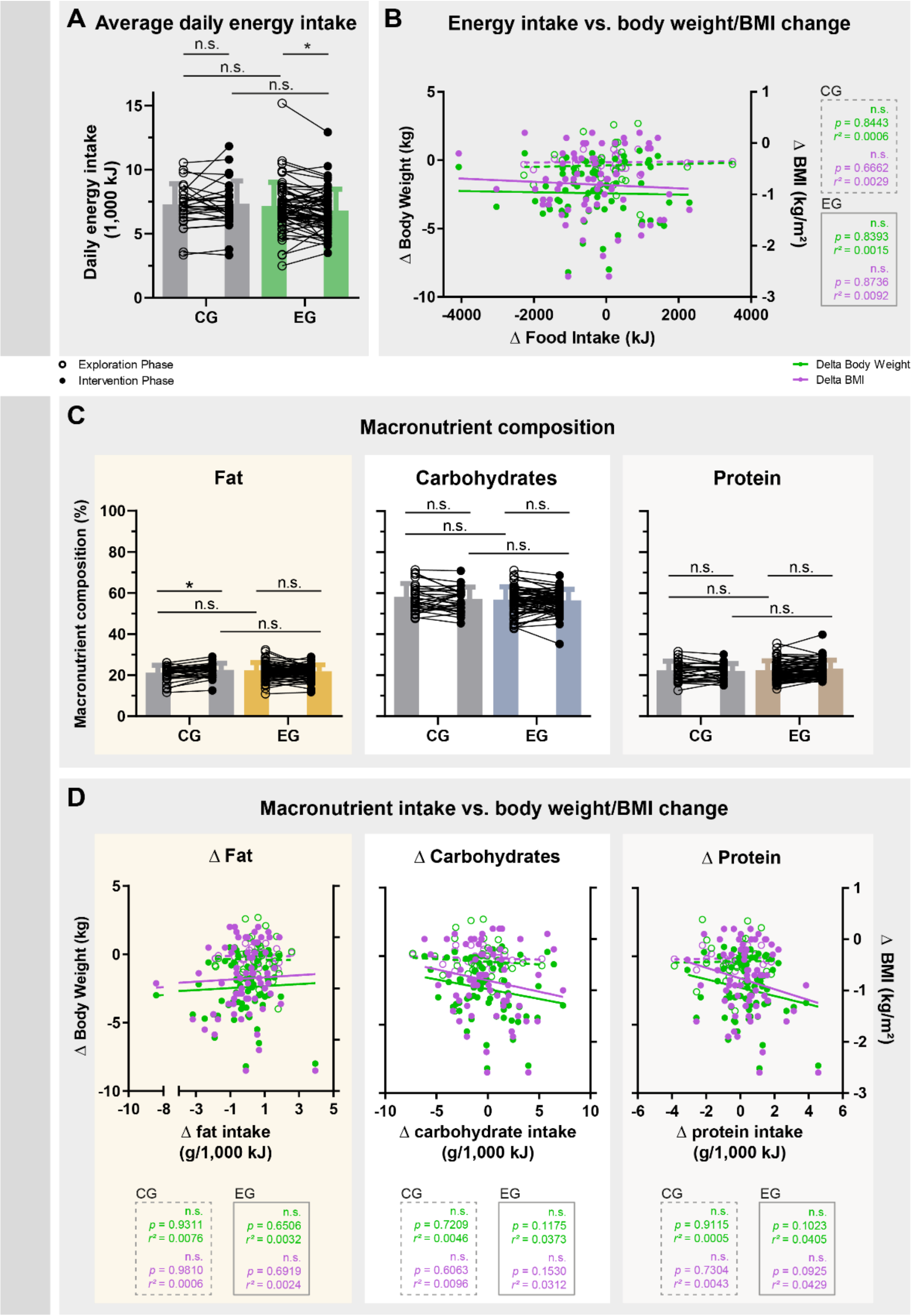


Fig. 3. Self-reported energy intake and macronutrient composition showed no consistent association with weight loss. (A) Average daily energy intake during the exploration and intervention phases in the control group (CG) and experimental group (EG). (B) Associations between changes in energy intake and changes in body weight or BMI. (C) Macronutrient composition during exploration and intervention, shown as percentage of total intake from fat, carbohydrates, and protein. (D) Associations between changes in macronutrient intake and changes in body weight or BMI. Statistical annotations indicate repeated-measures ANOVA or mixed-effects models with Bonferroni correction and linear regression analyses where applicable. Adjusted between-group analyses are reported in the main text and tables.

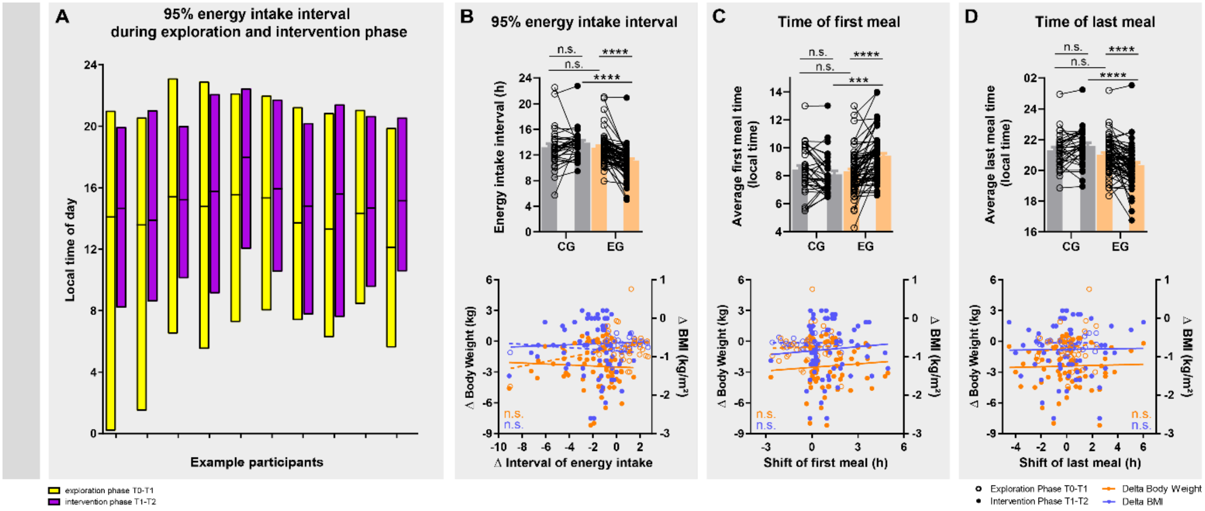


Fig. 4. Additional meal timing characteristics and their association with weight loss. (A) Daily 95% caloric intake interval in 10 example participants during the exploration and intervention phases. Bars indicate the time window containing 95% of caloric intake events. (B) Changes in eating interval duration and their associations with changes in body weight or BMI. (C, D) Changes in the timing of the first (C) and last (D) caloric intake event and their associations with changes in body weight or BMI. Statistical annotations indicate repeated-measures ANOVA or mixed-effects models with Bonferroni correction and linear regression analyses where applicable. Adjusted between-group analyses are reported in the main text and tables.

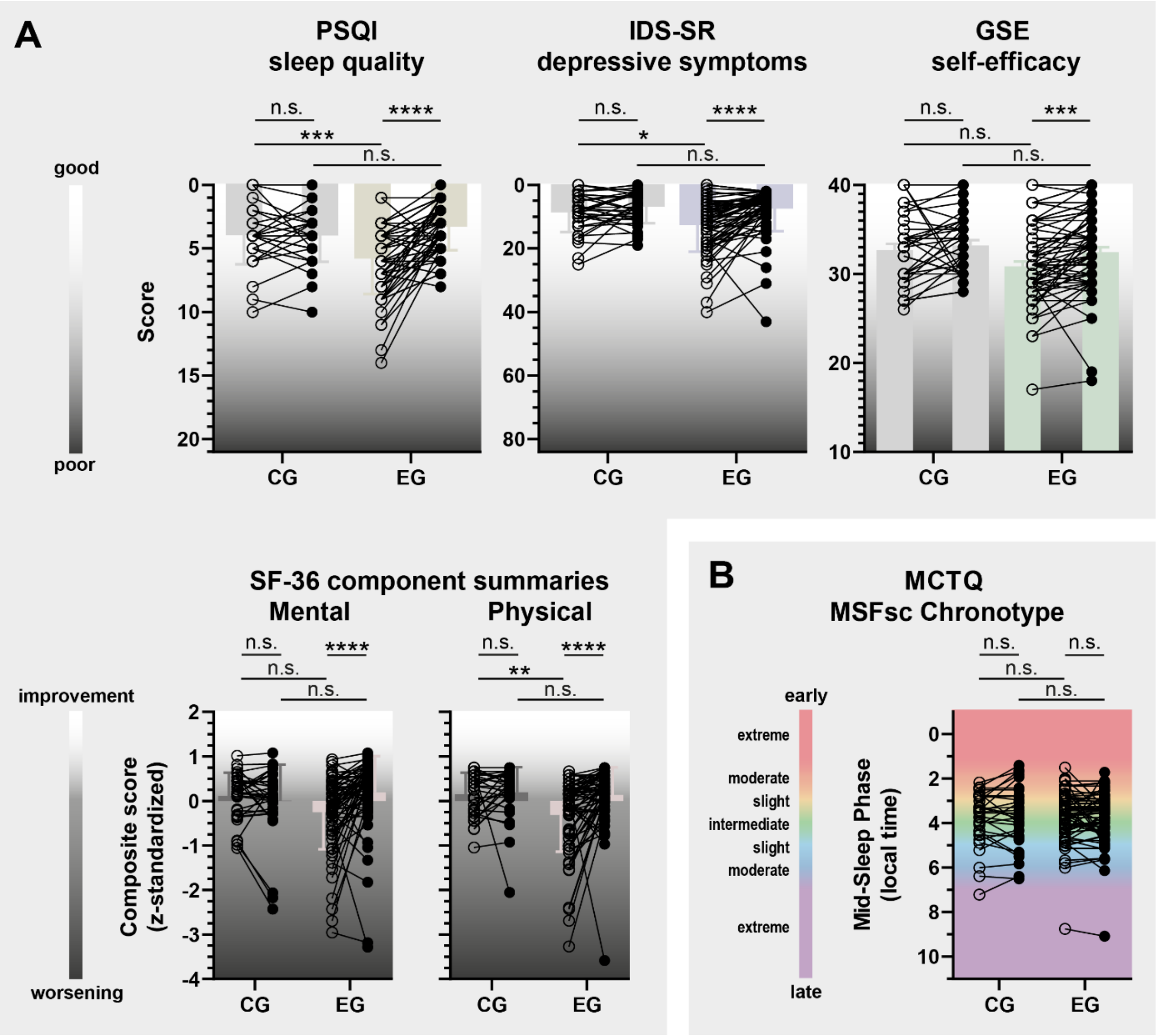


Fig. 5. Changes in self-reported well-being and chronotype during the intervention. (A) Sleep quality (PSQI), depressive symptoms (IDS-SR), self-efficacy (GSE), and SF-36 mental and physical component summary scores (MCS and PCS) in the control group (CG, gray) and experimental group (EG, colored). Lower PSQI and IDS-SR scores and higher SWE and SF-36 scores indicate improvement. SF-36 component scores were derived from z-standardized subscale scores. (B) Chronotype, assessed as MSFsc, remained stable in both groups. Statistical annotations indicate repeated-measures ANOVA or mixed-effects models with Bonferroni correction. Adjusted between-group analyses are reported in Table 2.

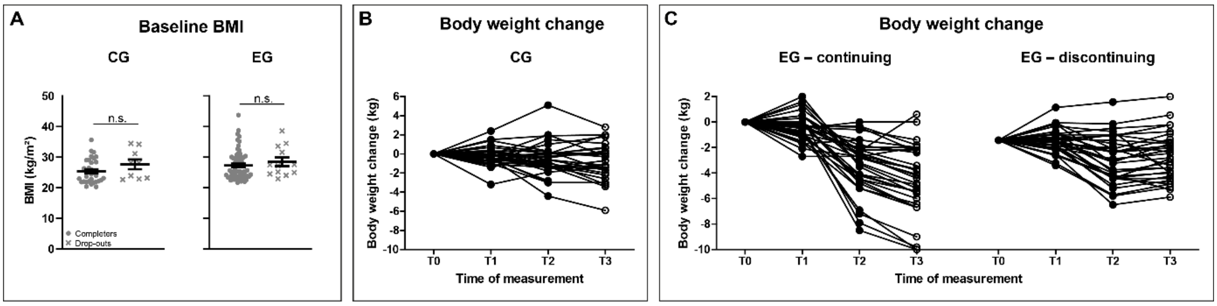


Fig. S1. Baseline BMI and individual body-weight trajectories. (A) Baseline BMI in completers and drop-outs by group. (B) Individual body-weight trajectories in CG participants. (C) Individual body-weight trajectories in EG participants stratified by continuation or discontinuation of the intervention during follow-up. Body weight is shown as change from baseline.

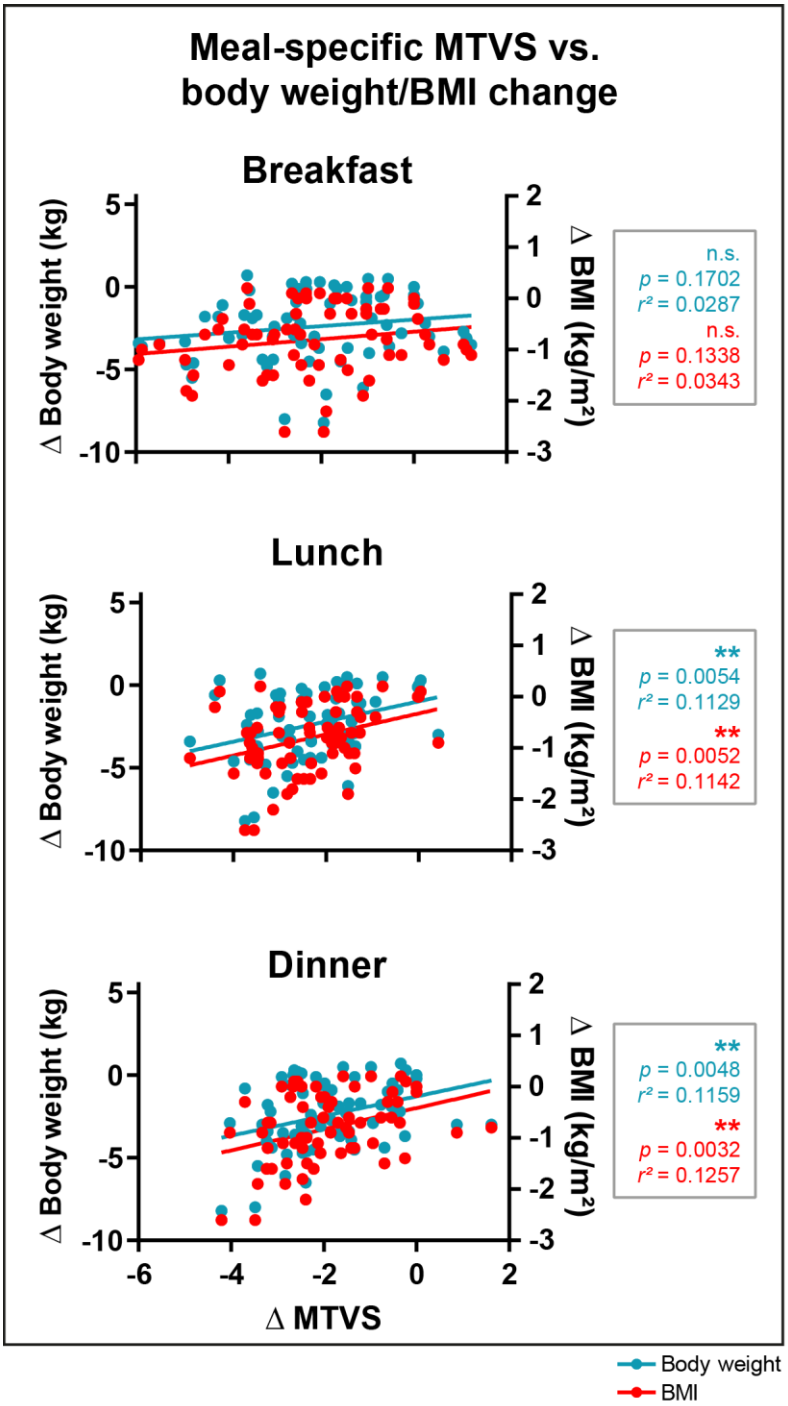


Fig. S2. Meal-specific regularity and body weight/BMI change. Associations between changes in mealtime variability score (MTVS) and changes in body weight or BMI for breakfast, lunch, and dinner in the experimental group. Lower MTVS values indicate greater meal regularity. Linear regression analyses indicated significant associations for lunch and dinner, but not breakfast. n = 67; $^{**}p \leq 0.01$.

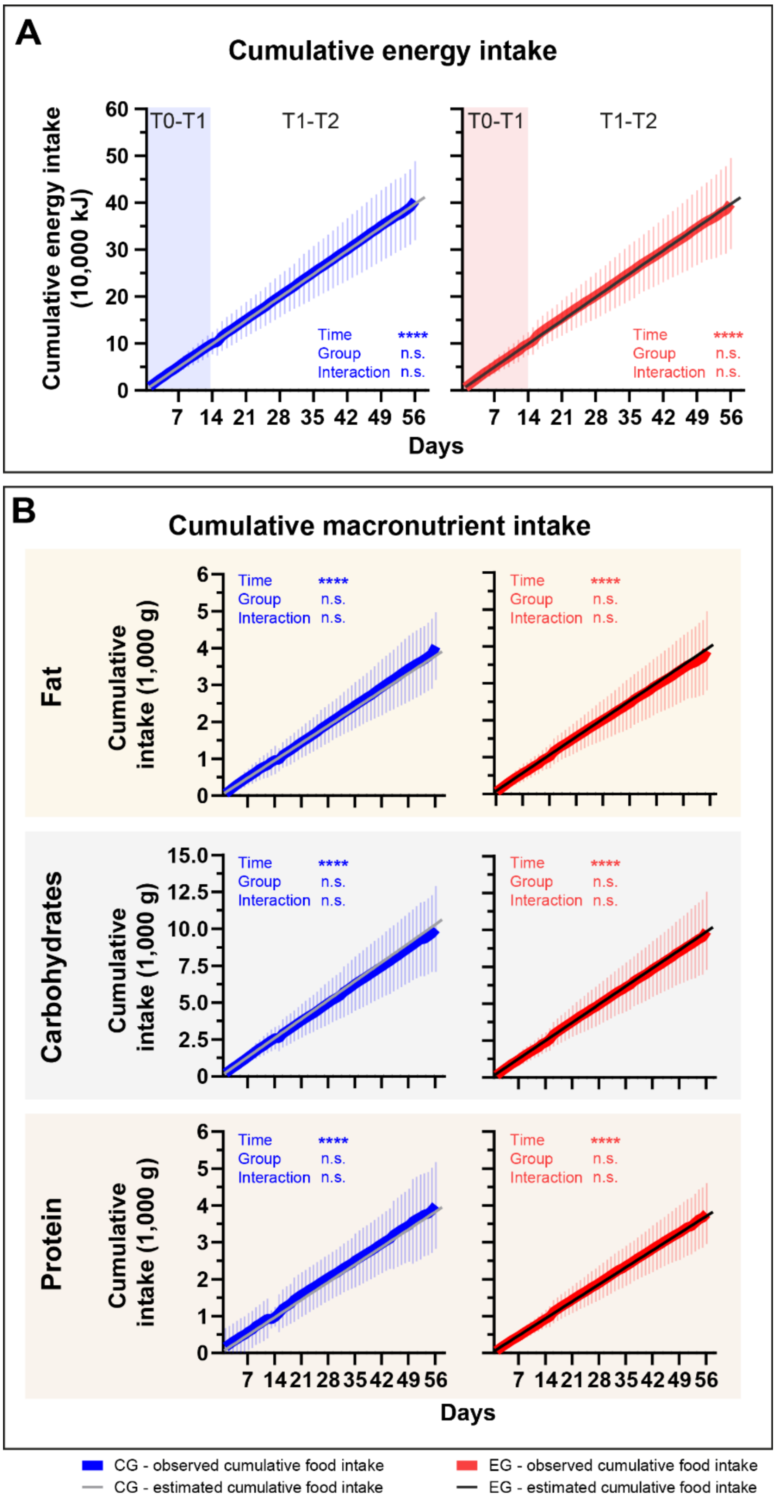


Fig. S3. Cumulative self-reported energy and macronutrient intake. (A) Observed cumulative energy intake in the control group (CG) and experimental group (EG) across the exploration (T0–T1) and intervention phases (T1–T2), compared with estimated cumulative intake based on extrapolation from average exploration-phase intake. (B) Observed and estimated cumulative macronutrient intake for fat, carbohydrates, and protein across study phases. Statistical annotations indicate repeated-measures ANOVA with Bonferroni correction. Estimated intake was calculated as a continuation of average intake during the exploration phase. Participants with implausible diary entries were excluded from the respective analyses.

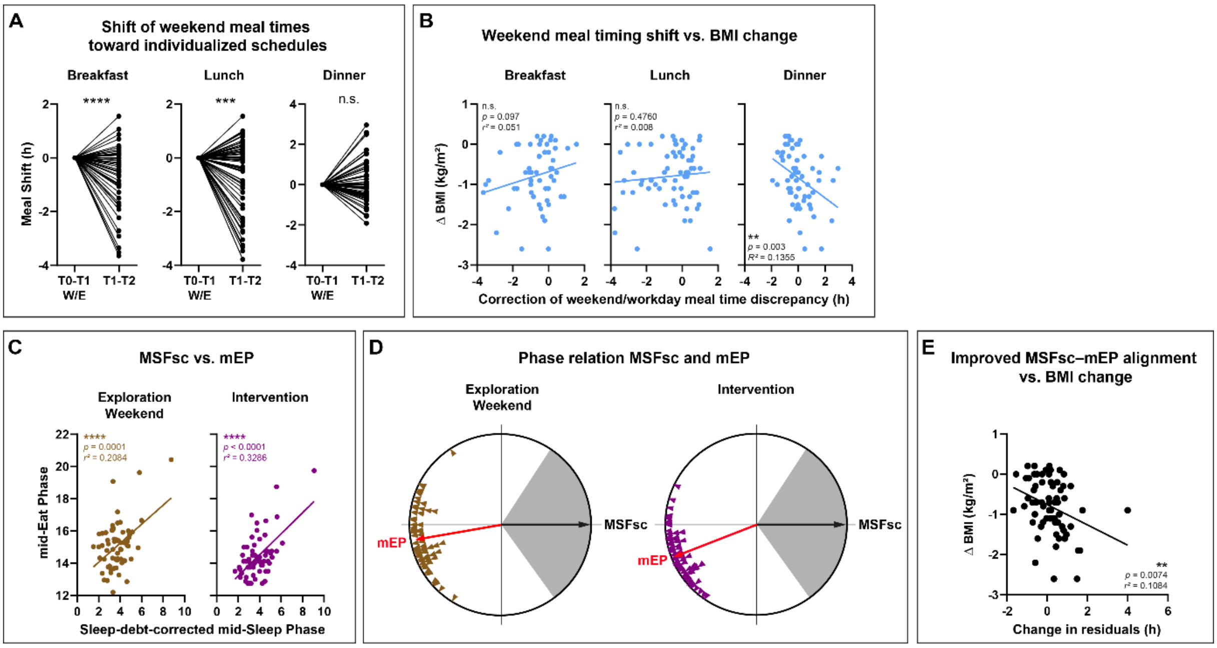


Fig. S4. Exploratory analyses of weekday–weekend meal timing and sleep–eating phase alignment. (A) Shifts in weekend meal timing toward calculated meal times for breakfast, lunch, and dinner during the intervention. (B) Associations between changes in weekday–weekend meal timing discrepancy and changes in BMI. (C) Associations between mid-sleep phase corrected for sleep debt (MSFsc) and mid-eat phase (mEP) during the exploration and intervention phases. (D) Circular representation of the phase relationship between MSFsc and mEP during exploration and intervention. (E) Association between changes in MSFsc –mEP alignment, quantified by residuals from the MSFsc –mEP relationship, and changes in BMI. Negative BMI changes indicate BMI reduction. Statistical annotations indicate paired tests or linear regression analyses, as appropriate.

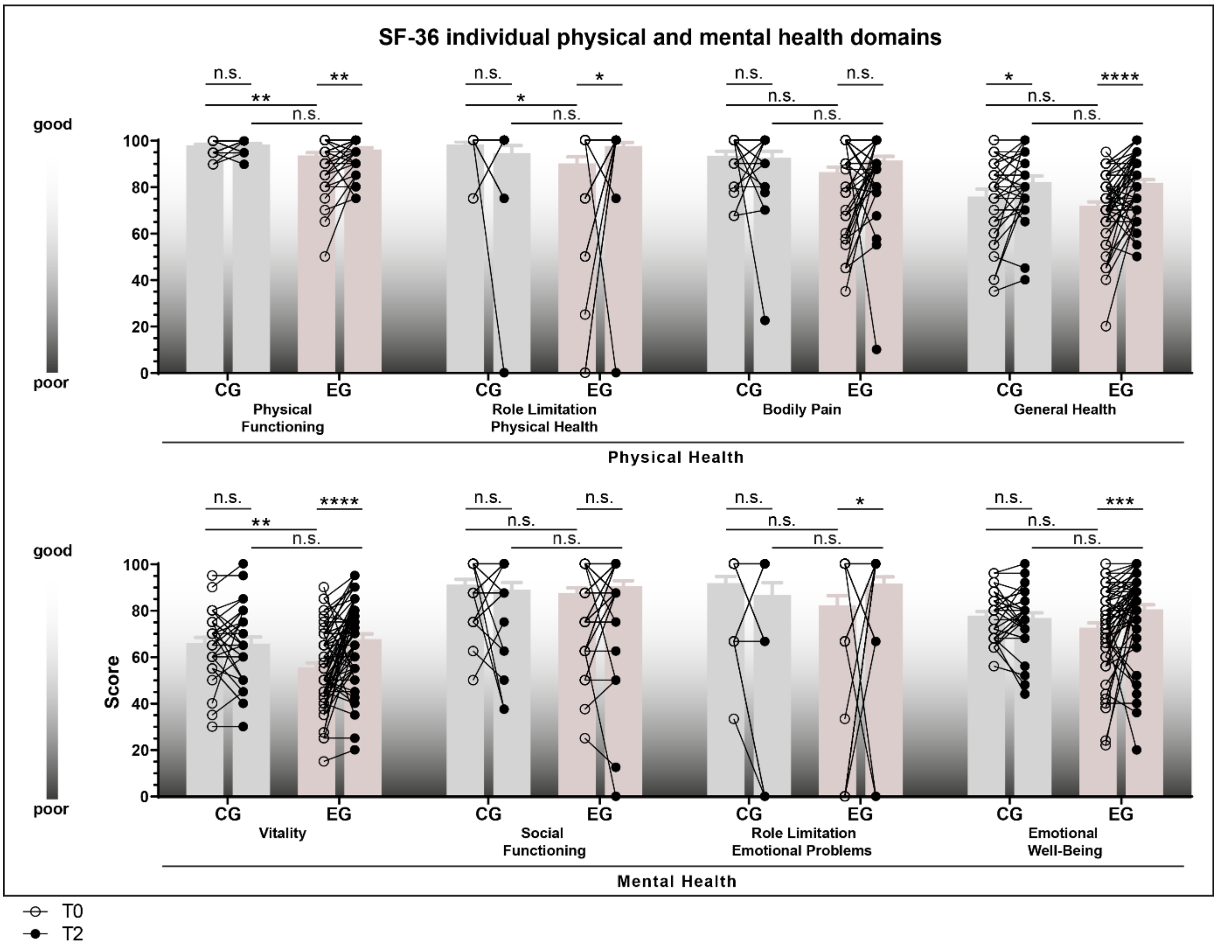


Fig. S5. Changes in individual SF-36 subscale scores during the intervention. SF-36 domains are shown for the control group (CG, gray) and experimental group (EG, colored) at baseline (T0) and after the intervention (T2), including physical functioning (PF), role limitations due to physical health (RP), bodily pain (BP), general health (GH), vitality (VT), social functioning (SF), role limitations due to emotional problems (RE), and emotional well-being/mental health (MH). Higher scores indicate better health status. Statistical annotations indicate repeated-measures ANOVA or mixed-effects models with Bonferroni correction, as appropriate. Sample sizes may vary due to missing questionnaire data.
