## Supplemental Tab. S1 for "Time to Eat: Effects of Personalized Regular Meal Schedules on Body Weight and Mental and Physical Well-Being—A Randomized Controlled Pilot Trial"

**Table S1. Baseline characteristics of completers included in the primary analysis.**

| Characteristic | CG | EG | p value |
|---|---|---|---|
| n | 33 | 67 | — |
| Sex, male/female | 8/25 | 26/41 | 0.1814 |
| Age, years | 32.64 ± 11.81 (20.00–60.00) | 38.18 ± 14.41 (19.00–64.00) | 0.0440 |
| BMI at baseline (T0), $kg/m^2$ | 25.46 ± 3.83 (20.40–35.76) | 27.39 ± 4.62 (22.02–43.60) | 0.0305 |
| Body weight at baseline (T0), kg | 75.04 ± 16.94 (54.20–134.60) | 82.73 ± 16.72 (58.50–129.00) | 0.0359 |
| MTVS during exploration | 3.35 ± 0.81 (1.32–4.79) | 3.89 ± 0.90 (1.11–5.76) | 0.0034 |
| Energy intake during exploration, kJ/day | 7,211.47 ± 1,596.27 (3,349.93–10,537.38) | 7,070.77 ± 1,854.11 (2,505.08–15,184.57) | 0.7043 |
| Fat intake during exploration, g/1,000 kJ | 9.29 ± 1.39 (5.40–11.28) | 9.67 ± 1.88 (4.70–19.08) | 0.2671 |
| Carbohydrate intake during exploration, g/1,000 kJ | 26.05 ± 4.08 (19.99–34.45) | 24.99 ± 3.88 (16.08–36.55) | 0.2380 |
| Protein intake during exploration, g/1,000 kJ | 9.77 ± 1.85 (5.93–13.70) | 9.70 ± 2.09 (6.03–16.78) | 0.8648 |
| Eating interval during exploration, h | 13.29 ± 2.91 (8.50–22.53) | 13.00 ± 2.26 (7.93–21.05) | 0.6230 |
| First meal time during exploration, h | 8.39 ± 1.68 (5.50–13.00) | 8.24 ± 1.52 (4.28–13.00) | 0.6633 |
| Last meal time during exploration, h | 21.26 ± 1.14 (18.85–24.95) | 20.96 ± 1.14 (18.33–25.20) | 0.2198 |

**Table note:** Values are mean ± SD (range) unless otherwise indicated. Baseline refers to T0 for age, BMI, and body weight and to the exploration phase for eating-behavior variables. Between-group comparisons were performed using Welch's t-tests for continuous variables and Fisher's exact test for sex. Sample sizes for dietary and meal timing variables may vary due to missing or excluded dietary records. MTVS, mealtime variability score.
