## Supplemental Tab. S2 for "Time to Eat: Effects of Personalized Regular Meal Schedules on Body Weight and Mental and Physical Well-Being—A Randomized Controlled Pilot Trial"

**Supplementary Table S2. Exploratory multivariable regression model for BMI change in the experimental group**

| Predictor | Estimate | SE | 95% CI | t | p value | VIF |
|---|---|---|---|---|---|---|
| Intercept | −0.1691 | 0.2434 | −0.6594 to 0.3212 | 0.6947 | 0.4908 | — |
| Δ MTVS | 0.2582 | 0.1056 | 0.0455 to 0.4709 | 2.445 | 0.0185 * | 1.164 |
| Δ energy intake | 0.0000278 | 0.0000711 | −0.000115 to 0.000171 | 0.3917 | 0.6971 | 1.048 |
| Δ eating interval | −0.0461 | 0.0551 | −0.1571 to 0.0650 | 0.8355 | 0.4079 | 1.475 |
| Δ mSP–mEP residuals | −0.2217 | 0.1233 | −0.4700 to 0.0267 | 1.798 | 0.0789 | 1.116 |
| Δ breakfast weekday–weekend discrepancy | 0.1457 | 0.0840 | −0.0234 to 0.3148 | 1.736 | 0.0895 | 1.247 |
| Δ dinner weekday–weekend discrepancy | −0.3071 | 0.0851 | −0.4786 to −0.1357 | 3.608 | 0.0008 *** | 1.079 |

Model summary: n = 52; $R^2$ = 0.3970; adjusted $R^2$ = 0.3167; F(6,45) = 4.939, p = 0.0006.

Table note: Dependent variable was Δ BMI. Negative Δ BMI values indicate BMI reduction. The model included experimental-group participants with complete data for all predictors. Δ MTVS reflects change in mealtime variability score, with lower MTVS values indicating greater meal regularity. mSP–mEP residuals quantify individual deviation between mid-sleep phase and mid-eat phase. Variance inflation factor (VIF) values indicated no relevant multicollinearity. Residuals did not significantly deviate from normality based on Shapiro–Wilk testing.
