## Supplemental Tab. S3 for "Time to Eat: Effects of Personalized Regular Meal Schedules on Body Weight and Mental and Physical Well-Being—A Randomized Controlled Pilot Trial"

**Table S3.** Questionnaires collected at T0 and T2 to assess well-being and chronotype.

| Construct | Questionnaire | Components / domains assessed |
|---|---|---|
| Sleep quality | Pittsburgh Sleep Quality Index (PSQI)[1] | Subjective sleep quality, sleep latency, sleep duration, habitual sleep efficiency, sleep disturbances, use of sleep medication, and daytime dysfunction. |
| Health-related quality of life | 36-Item Short Form Health Survey (SF-36)[2] | Physical functioning, role limitations due to physical health, bodily pain, general health, vitality, social functioning, role limitations due to emotional problems, and mental health. |
| Self-efficacy | General Self-Efficacy Scale (GSE) [3] | Optimistic self-beliefs regarding coping with difficult demands, independent problem solving, and attribution of success to one's own competence. |
| Depressive symptoms | Inventory of Depressive Symptomatology – Self-Report (IDS-SR)[4] | Depressive symptom severity across domains including mood, sleep, appetite and weight changes, concentration and decision making, self-esteem, future outlook, suicidality, energy, pleasure, psychomotor changes, anxiety/panic, somatic symptoms, and interpersonal sensitivity. |
| Chronotype and sleep timing | Munich Chronotype Questionnaire (MCTQ) [5] | Sleep timing on workdays and free days, including bedtimes, sleep onset, wake times, sleep duration, differences between workdays and free days, and derived chronotype measures such as mid-sleep phase on free days corrected for sleep debt (MSFsc). |
